# A robust coordinated molecular response following trauma promotes recovery and protects against chronic pain and neuropsychiatric symptoms

**DOI:** 10.64898/2026.11.07.26367466

**Authors:** Brittanie L Winfield, Samuel A McLean, Erica M Branham, Natalie Stanley, Stacey L House, Francesca L Beaudoin, Jennifer S Stevens, Thomas C Neylan, Gari D Clifford, Laura T Germine, John P Haran, Alan B Storrow, Christopher Lewandowski, Paul I Musey, Phyllis L Hendry, Sophia Sheikh, Ronald C Kessler, Kerry J Ressler, Sarah D Linnstaedt

**Affiliations:** Institute for Trauma Recovery, University of North Carolina at Chapel Hill, Chapel Hill, NC, USA; Department of Anesthesiology, University of North Carolina at Chapel Hill, Chapel Hill, NC, USA; Curriculum in Bioinformatics and Computational Biology, University of North Carolina at Chapel Hill, Chapel Hill, NC, USA; Department of Emergency Medicine, University of North Carolina at Chapel Hill, Chapel Hill, NC, USA; Department of Psychiatry, University of North Carolina at Chapel Hill, Chapel Hill, NC, USA; Curriculum in Genetics and Molecular Biology, University of North Carolina at Chapel Hill, Chapel Hill, NC, USA; Department of Computer Science, University of North Carolina at Chapel Hill, Chapel Hill, NC, USA; Department of Genetics, University of North Carolina at Chapel Hill, Chapel Hill, NC, USA; Computational Medicine Program, University of North Carolina at Chapel Hill, Chapel Hill, NC, USA; Department of Emergency Medicine, Washington University School of Medicine, St. Louis, MO, USA; Department of Epidemiology, Brown University, Providence, RI, USA; Department of Emergency Medicine, Brown University, Providence, RI, USA; Department of Psychiatry and Behavioral Sciences, Emory University School of Medicine, Atlanta, GA, USA; Departments of Psychiatry and Neurology, University of California San Francisco, San Francisco, CA, USA; Department of Biomedical Engineering, Johns Hopkins University, Baltimore, MD, USA; Department of Anesthesia and Critical Care Medicine, Johns Hopkins University, Baltimore, MD, USA; Department of Neurology, Albert Einstein College of Medicine, New York, NY, USA; Department of Emergency Medicine, University of Massachusetts Chan Medical School, Worcester, MA, USA; Department of Emergency Medicine, Vanderbilt University Medical Center, Nashville, TN, USA; Department of Emergency Medicine, Henry Ford Health System, Detroit, MI, USA; Department of Emergency Medicine, Indiana University School of Medicine, Indianapolis, IN, USA; Department of Emergency Medicine, University of Florida College of Medicine -Jacksonville, Jacksonville, FL, USA; Department of Health Care Policy, Harvard Medical School, Boston, MA, USA; Division of Depression and Anxiety, McLean Hospital, Belmont, MA, USA; Department of Psychiatry, Harvard Medical School, Boston, MA, USA

**Author notes:** <u>Manuscript correspondence:</u> Sarah D. Linnstaedt, PhD, University of North Carolina at Chapel Hill, Campus Box #7010, Chapel Hill, NC 27599-7010. **Author Contributions** B.L.W., S.A.M., and S.D.L. conceived and designed the study and contributed to hypothesis development and biological interpretation of the findings. B.L.W. performed all data analyses, generated all figures and tables, and prepared the initial manuscript draft. B.L.W. and S.D.L. worked collaboratively to further develop and revise the manuscript. S.D.L. supervised the study, guided the analytical strategy, led overall biospecimen collection, storage, and processing, and provided scientific oversight. S.A.M. led the parent AURORA study, overseeing its design and implementation, including coordination of participant enrollment, longitudinal follow-up, and study operations. E.M.B. led processing of the steroid hormone assay data and contributed to figure design. N.S. provided methodological review of the machine learning approaches. S.A.M. and E.M.B. critically reviewed and revised the manuscript. The remaining authors are members of the AURORA Study Group who contributed to the design and implementation of the parent AURORA study, including study conceptualization, protocol development, selection of study measures, participant recruitment, data collection, and/or study oversight. All authors revie wed and approved the final manuscript.

## Abstract

The biological mechanisms governing recovery versus chronicity of adverse posttraumatic neuropsychiatric sequelae (APNS) remain poorly understood. Here, we characterized longitudinal molecular changes among trauma survivors across ten APNS domains, including pain, anxiety, and depression. Recovery across these diverse clinical outcomes was associated with a shared, temporally coordinated transcriptional signature, characterized by early innate immune activation, subsequent homeostatic and wound-healing processes, and later adaptive immune responses. In contrast, chronic APNS trajectories exhibited limited transcriptional remodeling. We derived and externally validated an RNA-based Recovery Score (RRS) capturing the shared molecular response underlying recovery. Higher RRS was associated with greater clinical recovery, elevated peritraumatic cortisol, and lower cumulative stress exposure. Drug-response profiling identified candidate compounds predicted to mimic recovery-associated transcriptional changes. Together, these findings support posttraumatic recovery as an active biological process and identify shared molecular pathways that may be harnessed to prevent multiple adverse posttraumatic outcomes.

## Main

Traumatic stress exposures (TSEs) are a nearly universal human experience and include assaults, military combat, natural disasters, motor vehicle collisions and other life-threatening experiences. Although most individuals recover following TSEs, a substantial subset develop adverse posttraumatic neuropsychiatric sequelae (APNS), such as posttraumatic stress, chronic pain, depressive symptoms, anxiety, sleep disturbance, and cognitive impairment.^1,2^ Collectively, these APNS contribute substantially to disability, diminished quality of life, increased comorbidity and suicide risk, and considerable societal burden worldwide.^3,4^ Traditionally, APNS have been investigated as distinct clinical entities. However, growing evidence demonstrates that these outcomes frequently co-occur^2^ and may arise, at least in part, from shared molecular processes initiated in response to TSE.^5,6^ Defining shared molecular programs that promote recovery across APNS could reveal therapeutic targets with the potential to improve multiple trauma-related outcomes simultaneously.

Although substantial research has focused on the biological mechanisms contributing to APNS chronicity, comparatively less attention has been directed toward understanding recovery as an active biological process. One approach to addressing this gap is to identify molecular processes that differentiate individuals who recover from those who develop chronic APNS.^7,8^ The potential value of this approach is supported by evidence that recovery following TSE involves coordinated biological responses^9-13^ (e.g., glucocorticoid signaling, inflammatory pathways) rather than isolated molecular events. However, whether recovery involves a shared, coordinated molecular program across diverse APNS remains unclear. Moreover, most prior studies have examined biological signals at single timepoints after TSE, and/or have focused on individual APNS, rather than evaluating how molecular processes dynamically evolve from the immediate aftermath of TSE through longer-term recovery. Characterizing these dynamic molecular programs beginning in the hours after TSE is particularly important, as available evidence suggests that the initial biological response may shape subsequent trajectories of recovery and chronicity.^14,15^

In this study, we leveraged longitudinal molecular and APNS outcome data from trauma survivors enrolled in the AURORA cohort to identify shared molecular processes associated with recovery versus chronicity across ten APNS domains. AURORA enrolled participants in the emergency department (ED) within 72 hours of TSE and assessed ten APNS outcomes over six months post-TSE.^1^ By integrating longitudinal transcriptomic data with inferred transcription factor activity, estimated immune cell fractions, circulating cytokines, and peritraumatic steroid hormone measurements, we characterized molecular programs associated with recovery, developed an individualized RNA-based Recovery Scores (RRS) to quantify recovery-associated transcriptional remodeling, identified biological factors associated with the RRS, and used drug-response signature analysis to identify compounds predicted to mimic recovery-associated transcriptional changes.

## Results

### Cohort description and study design

We utilized samples and data from AURORA, a longitudinal observational study of individuals enrolled in the ED within 72 hours of TSE.^1^ For this nested cohort study, paired blood samples collected at enrollment and six months (6M) post-TSE were available from 245 participants who were representative of the full AURORA cohort (**Ext. Data Fig 1a**; **Supplementary Table 1**). Most participants were enrolled following motor vehicle collisions (MVCs) and were Black, female, socioeconomically disadvantaged, with high levels of peritraumatic distress (**Ext. Data Fig 1b**). Previously characterized latent class trajectories described longitudinal symptom patterns across ten APNS domains, including pain, depression, hyperarousal, mental fatigue, and sleep disturbance (**Ext. Data Fig 1c**).^2,16^ For each APNS domain, we identified the trajectories characterized by the greatest recovery (“Recovery”) and the greatest chronicity (“Chronic”) (**Ext. Data Fig 1d**). We examined longitudinal RNA expression changes between the ED and 6M timepoints across APNS trajectories, including comparisons between Recovery and Chronic trajectories (**Ext. Data Fig 1e**). We then characterized associated biological pathways, transcription factor activity, immune cell fractions, and cytokines (**Ext. Data Fig 1f-h**), and identified candidate pharmacologic perturbations using the Connectivity Map^17^ (LINCS L1000^18^). We also developed and validated an RNA Recovery Score (RRS) and examined its associations with pre- and peritraumatic factors (**Ext. Data Fig 1i-j**). Finally, in a subsample with an additional 2-week (2WK) post-TSE timepoint (n=40), we further characterized the timing of recovery-associated molecular changes (**Ext. Data Fig 1k**).

### Recovery following TSE is characterized by extensive longitudinal transcriptomic changes

To identify molecular processes associated with recovery following TSE, we first examined longitudinal changes in gene expression between ED and 6M timepoints across trajectories for each of the ten APNS domains (**Fig 1**). Across all ten domains, Recovery trajectories exhibited extensive gene expression changes with 4,360-7,319 of 12,513 tested transcripts (35-58%) differentially expressed between ED and 6M (FDR<0.01), **Fig 1a-j**; teal outlined volcano plots). In contrast, Chronic trajectories consistently exhibited substantially fewer longitudinal gene expression changes (**Fig 1a-j**; burgundy outlined volcano plots). To account for differences in trajectory sample sizes, we performed sensitivity analyses involving repeated downsampling to 20 participants per trajectory. Recovery trajectories continued to exhibit substantially more differentially expressed transcripts than Chronic trajectories across all ten APNS domains (**Ext. Data Fig 2**). Given the frequent co-occurrence of APNS following TSE, both in prior literature (e.g.^19-22^) and within AURORA (**Ext. Data Fig 3**), we next examined the extent to which recovery-associated transcriptional changes were shared across APNS domains. Strikingly, 3,574 genes exhibited concordant longitudinal expression changes across all Recovery trajectories, including 1,829 with higher expression at the ED and 1,745 with higher expression at 6M (**Fig. 1a-j**; **Supplementary Tables 2-3**). These findings identify a shared longitudinal transcriptional program associated with recovery across diverse posttraumatic outcomes.

**Fig 1.**
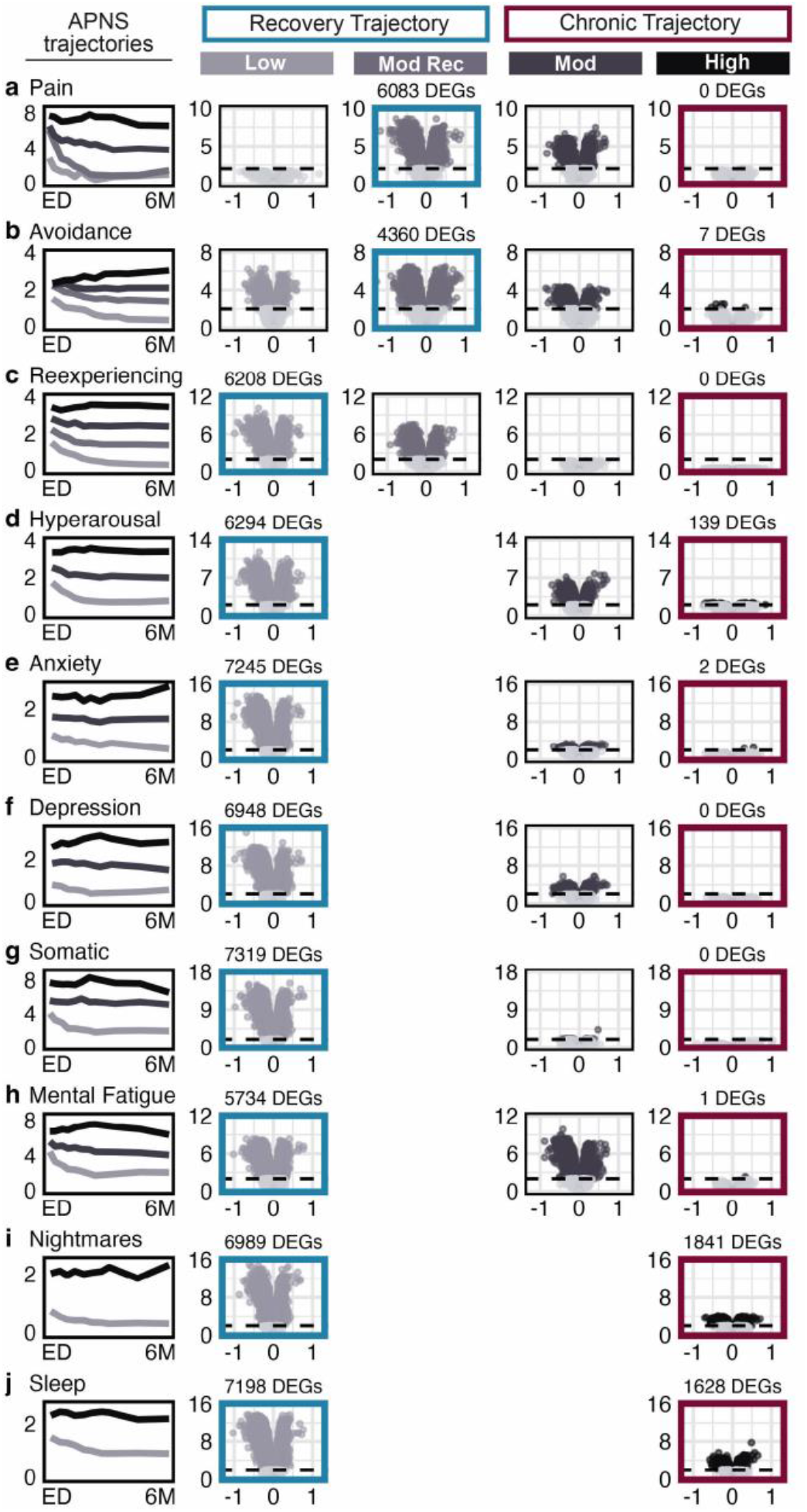
Across APNS, trajectories with the greatest symptom resolution following traumatic stress exposure (TSE) show widespread longitudinal transcriptional changes, while those with persistent symptoms show minimal changes. Participants were assigned to post-TSE symptom trajectories for each of ten adverse posttraumatic neuropsychiatric sequelae (APNS) using growth mixture modeling of longitudinal self-reported symptom severity (2-4 trajectories per APNS). For each trajectory, paired differential expression analyses compared RNA profiles from the initial emergency department (ED) visit to the 6-month (6M) follow-up. Panels show **a.** pain, **b.** avoidance, **c.** reexperiencing, **d.** hyperarousal, **e.** anxiety, **f.** depression, **g.** somatic symptoms, **h.** mental fatigue, **i.** nightmares, and **j.** sleep disturbance. Longitudinal gene expression changes were greatest in Recovery trajectories (blue outlines) and smallest in Chronic trajectories (red outlines). In the volcano plots, the x-axis indicates log2 fold change (6M versus ED), and the y-axis indicates -log10(p value); the dashed line denotes the significance threshold (FDR-adjusted p = 0.01), and points represent individual genes, colored if statistically significant and gray otherwise.

### Molecular processes characterizing recovery include robust peritraumatic neutrophil and IL-6 responses and a coordinated immune response over time

We next examined the 3,574 shared recovery-associated differentially expressed genes (DEGs) to characterize the biological processes underlying longitudinal recovery. Across recovery trajectories, longitudinal gene expression patterns revealed an early predominance of innate immune and inflammatory pathways followed by a shift toward cellular maintenance and adaptive immune processes (**Fig 2a-e**). Transcripts with higher expression at the ED visit were enriched for neutrophil and innate immune functions (**Fig 2a**, **blue circles**), consistent with Gene Ontology Biological Process (GO-BP) and Reactome enrichment analyses. GO-BPs enriched at the ED in the Recovery trajectory included neutrophil and granulocyte migration, myeloid leukocyte activation and migration, and regulation of innate and inflammatory immune responses (**Fig 2b, blue circles**). In contrast, fewer GO-BPs were enriched among transcripts with higher expression at 6M, primarily involving rRNA and tRNA metabolism, consistent with a transition toward cellular maintenance (**Fig 2b, teal circles**). Reactome analyses identified ED enrichment of neutrophil degranulation, Toll-like receptor signaling, interleukin signaling, and programmed cell death, while transcripts with higher expression at 6M were enriched for antigen receptor signaling, suggesting a shift toward adaptive immunity (**Supplementary Table 4**).

**Fig 2.**
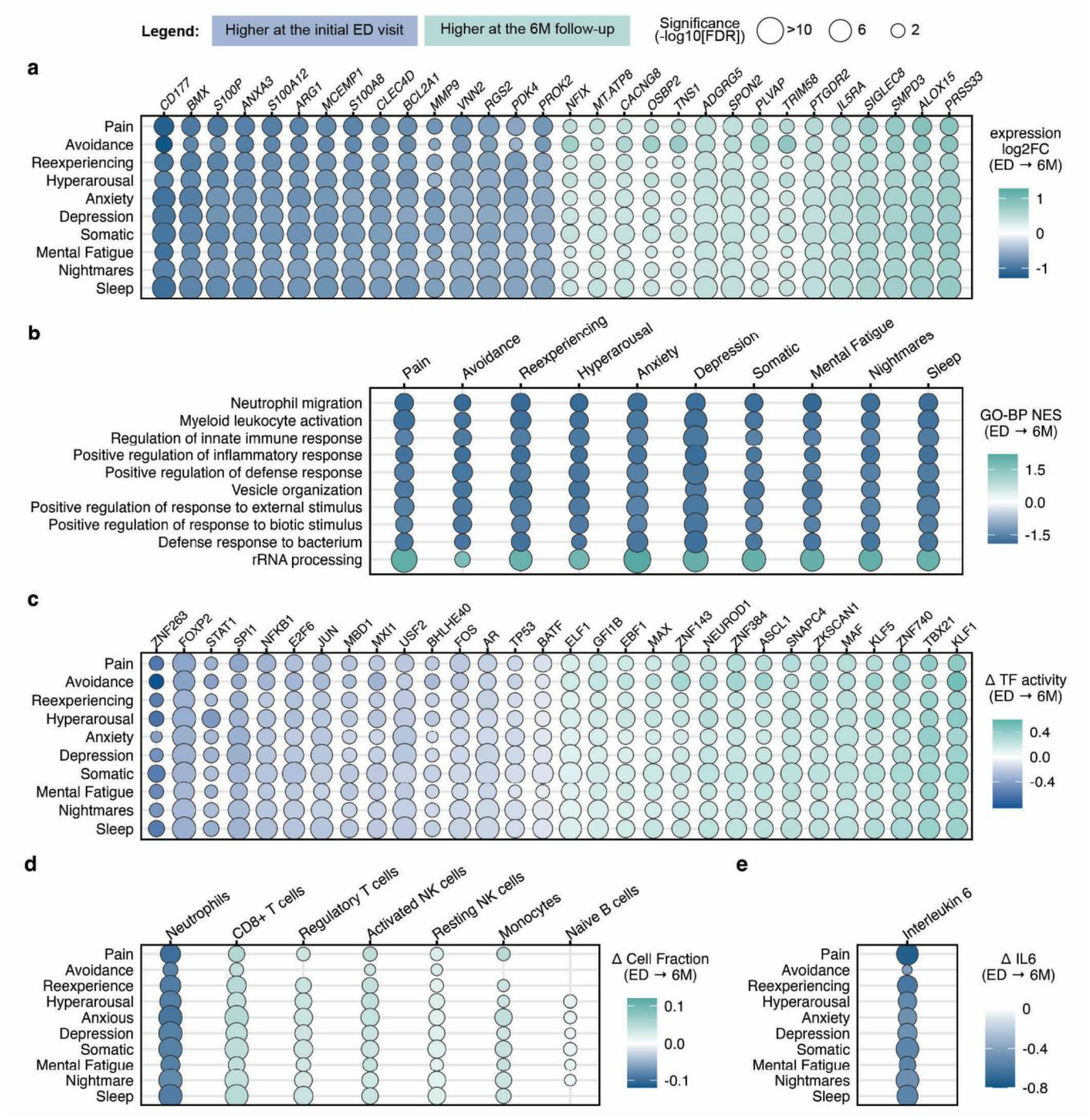
APNS symptom recovery following traumatic stress exposure (TSE) is associated with coordinated immune-related changes over time, including pronounced neutrophil and IL-6-mediated innate immune activation in the early aftermath of TSE. Panels a-e show Recovery trajectories across the ten APNS. Circle size reflects statistical significance (−log10 FDR); circle color reflects the direction and magnitude of change, with higher values at the initial emergency department (ED) visit in blue and higher values at the 6-month (6M) follow-up in teal. **a.** The 15 most upregulated differentially expressed genes (DEGs) at the ED and the 15 most upregulated DEGs at 6M, ranked by absolute log2 fold change. **b.** Gene Ontology Biological Process (GO-BP) enrichment among genes upregulated at the ED. **c.** The 15 most differentially active transcription factors (TFs) at the ED and the 15 most differentially active TFs at 6M, ranked by absolute change in activity. **d.** Longitudinal changes in estimated immune cell fractions, showing cell types with statistically significant changes (FDR < 0.01) in at least five APNS. **e.** Longitudinal changes in circulating IL-6 levels, the only cytokine among those tested (IL-6, IL-8, IL-10, TNF-α, and IFN-γ) showing a significant change consistently across APNS, with higher levels at the ED relative to 6M.

To determine whether these pathway-level changes reflected broader longitudinal changes in immune regulatory networks, we examined inferred transcription factor (TF) activity. Recovery trajectories exhibited coordinated longitudinal changes in TF activity across APNS, with an average of 82 differentially active TFs per APNS domain. TFs associated with inflammatory and myeloid biology^23^, including SPI1, NFKB1, JUN, FOS, and AR (androgen receptor), were consistently more active at the ED visit (**Fig 2c**, **blue circles**), whereas TFs associated with hematopoietic and immune-cell programming, including TBX21^24^ (T-bet), and MAF^25^, were more active at 6M (**Fig 2c**, **teal circles**). In contrast, Chronic trajectories exhibited comparatively limited changes in TF activity over time (**Supplementary Table 5**).

Longitudinal immune cell deconvolution analyses further supported coordinated changes in immune cell composition during recovery (**Fig 2d**). Across Recovery trajectories, estimated neutrophil abundance was elevated at the ED visit and declined significantly by 6M, whereas CD8+ T cells, regulatory T cells, activated natural killer (NK) cells and monocytes increased over time (**Supplementary Table 6**). Notably, 1,607 of the 3,574 shared recovery-associated genes (45%) were uniquely associated with neutrophil abundance, identifying neutrophils as the dominant immune cell correlate of early recovery-associated transcriptional changes (**Ext. Data Fig 4**). Consistent with these findings, circulating IL-6 concentrations were elevated at the ED visit and decreased over time, representing the most robust cytokine associated with recovery (**Fig 2e**; additional cytokines tested included IL-8, IL-10, TNF-α, IFN-γ; **Supplementary Table 7**). Together, these findings define a coordinated molecular recovery program characterized by the progressive resolution of an early neutrophil- and IL-6-associated innate immune response and an accompanying shift toward adaptive immune signaling over the subsequent six months.

### Drug-response profiling identifies candidate compounds predicted to mimic molecular recovery

Having identified coordinated transcriptional, immune cell, and cytokine changes associated with recovery, we next investigated whether pharmacologic perturbations could reproduce the recovery-associated transcriptional program. We queried Connectivity Map (CMap) using differential expression signatures derived from Recovery trajectories across multiple query sizes (**Ext. Data Fig 5**). These analyses identified compounds and mechanism of action (MOA) classes predicted to either mimic (**Fig 3a and c**) or oppose (**Fig 3b and d**) recovery-associated longitudinal transcriptional changes (**Supplementary Table 8)**.

**Fig 3.**
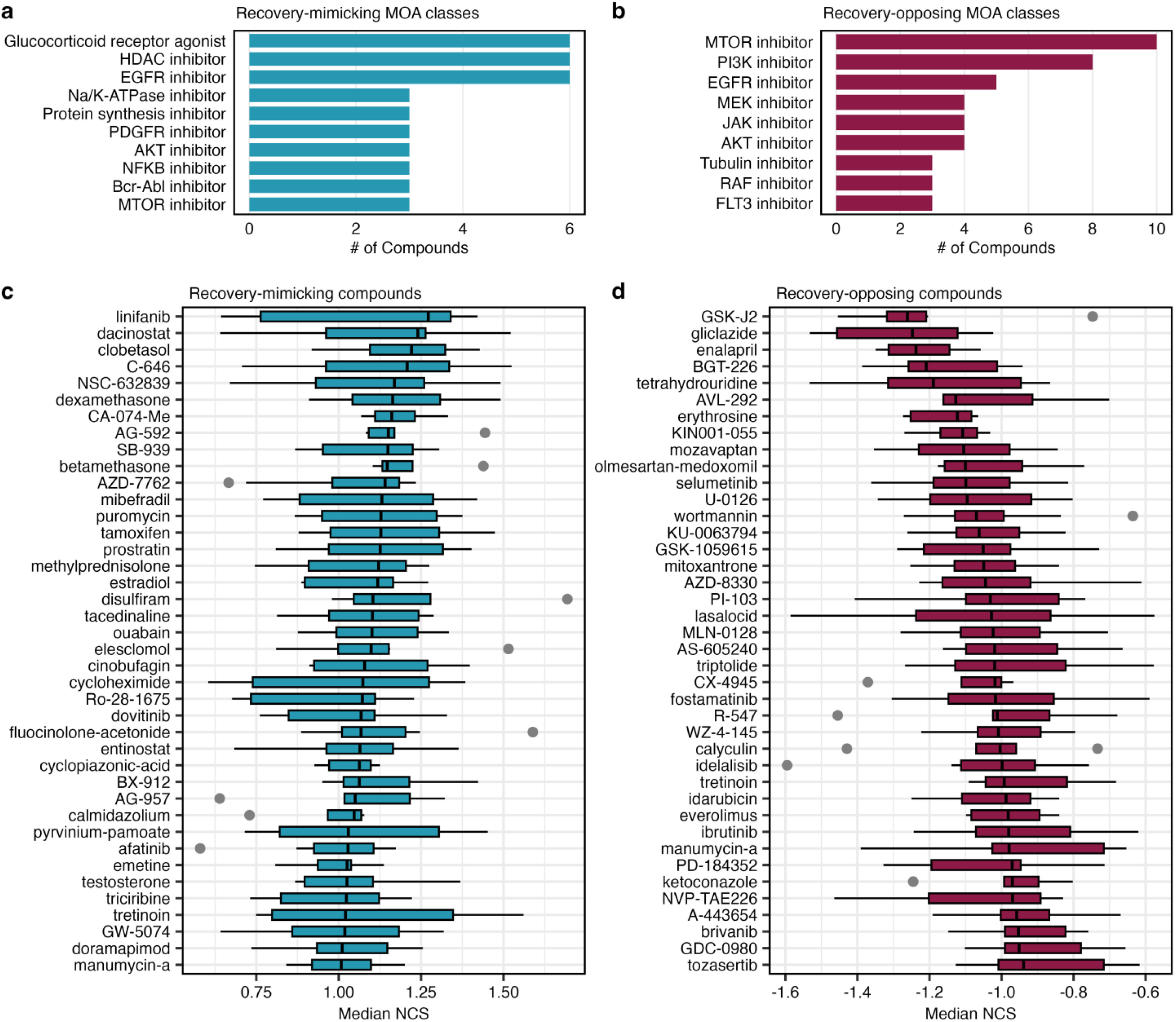
Drug perturbation analyses identify candidate compounds and mechanisms of action predicted to mimic recovery-associated transcriptional changes. Recovery-associated differentially expressed genes across three gene set sizes (top 50, 75, and 100 upregulated and downregulated) were submitted as CLUE queries against the LINCS Connectivity Map (Ext. Data Fig 5). Positive normalized connectivity score (NCS) values indicate perturbations predicted to mimic recovery-associated transcriptional patterns, and negative values indicate perturbations predicted to oppose these patterns. Compounds were classified as consistent mimickers or opposers based on their consistency across gene set sizes. Boxplots show the distribution of NCS values across matching LINCS signatures for each compound, reflecting variation across cell line, dose, and timepoint. **a.** Top mechanisms of action (MOA) compound classes among recovery-mimicking compounds. **b.** Top MOA compound classes among recovery-opposing compounds. **c.** Top 40 recovery-mimicking compounds ranked by median NCS. **d.** Top 40 recovery-opposing compounds ranked by median NCS.

Recovery-mimicking compounds, defined by transcriptional signatures positively associated with the recovery-associated expression program, converged on several shared MOAs. Glucocorticoid receptor agonists were among the strongest and most consistent recovery-mimicking classes and included dexamethasone and methylprednisolone (**Fig 3a and c**). Additional recovery-mimicking perturbations included protein kinase C (PKC) activators (e.g., prostratin) and the sex steroids estradiol and testosterone.

Conversely, recovery-opposing compounds, whose transcriptional signatures were negatively correlated with the recovery-associated expression program, were predominantly represented by inhibitors of mTOR and PI3K signaling (**Fig 3b and d).** Notably, several signaling pathways were represented among both recovery-mimicking and recovery-opposing perturbations; however, the specific compounds identified within each pathway were largely distinct. These findings identify pharmacologic pathways and compounds that may be leveraged to modulate shared molecular processes associated with recovery across multiple APNS domains.

### A recovery score derived from longitudinal gene expression data quantifies molecular recovery

To quantify the magnitude of the recovery-associated molecular response at the individual level, we used the 3,574 gene transcripts differentially expressed across all Recovery trajectories to derive an RNA Recovery Score (RRS) (**Fig 4a**, panels 1 and 2). Participants were classified according to recovery across the 10 APNS (**Ext. Data Fig 6a-b**) and randomly divided into training and held-out test cohorts, with an additional intermediate phenotype group retained as a separate comparison set. Elastic net modeling with bootstrap stability selection in the training cohort identified a stable 46-gene recovery signature from the shared recovery-associated transcriptome (**Ext. Data Fig 6c-d**), comprising 20 transcripts with higher expression at the ED visit and 26 with higher expression at 6M, combined into a weighted longitudinal expression score (RRS; **Fig 4a**, panels 3 and 4).

**Fig 4.**
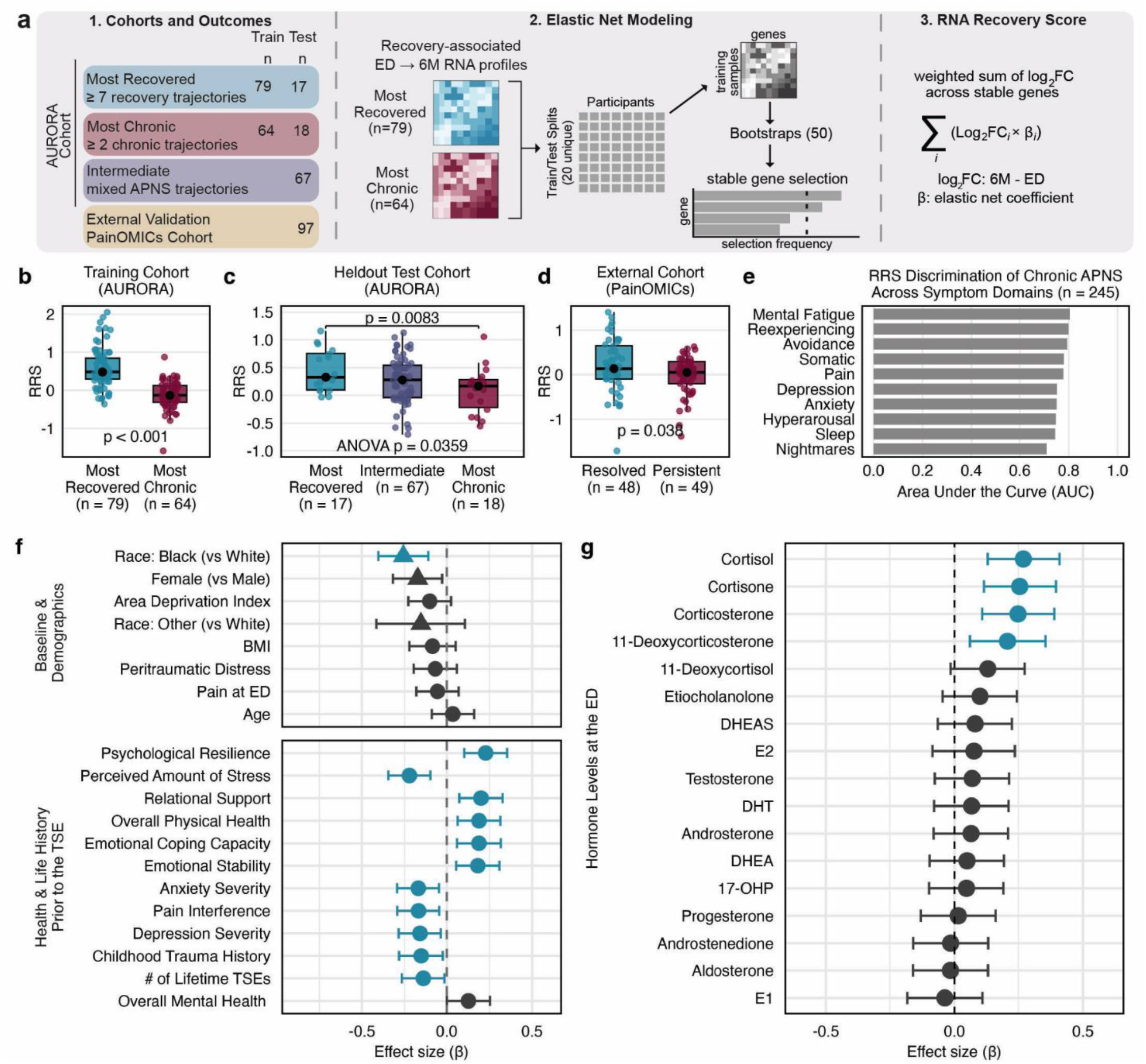
An RNA-based Recovery Score (RRS) derived from longitudinal transcriptional changes quantifies recovery-associated molecular changes and is positively associated with peritraumatic glucocorticoid levels and negatively associated with prior stress exposure. **a.** Construction of the RNA-based Recovery Score (RRS). **b.** RRS values in the AURORA training cohort were higher in the Most-Recovered group (t =9.69, p<0.001). **c.** In the held-out AURORA cohort, RRS values differed among Most-Recovered, Intermediate, and Most-Chronic participants (ANOVA F=3.44, p=0.036; Most-Recovered vs Most-Chronic t=2.83, p=0.008). **d.** In the external PainOMICs cohort, RRS values were higher in the Resolved group (t=2.11, p=0.038). **e.** AUC bar plots showing discrimination of Chronic versus non-Chronic trajectories across the ten APNS domains using the RRS. **f.** Associations between the RRS and baseline, demographic, clinical, and psychosocial, variables. Significant associations (FDR < 0.01) are shown in teal. **g.** Associations between the RRS and peritraumatic circulating steroid levels. Significant associations (FDR < 0.01) are shown in teal.

RRS values varied continuously, with higher scores reflecting greater alignment with the recovery-associated molecular response. In the training cohort, participants with greater clinical recovery exhibited higher RRS values (**Fig 4b**), and this association persisted, although attenuated, in the held-out test cohort (**Fig 4c**). RRS values in the intermediate phenotype group fell between these extremes (**Fig 4c**), consistent with the RRS as a continuous measure of molecular recovery. The RRS also differentiated Chronic versus non-Chronic trajectories across all ten APNS (**Fig 4e**). We further evaluated the RRS in the independent longitudinal PainOMICs RNA-sequencing cohort^26^. Of the 46 genes comprising the score, 43 were present in the PainOMICs dataset, where higher RRS values were associated with pain recovery (**Fig 4d**). Participant-level RRS values are provided in **Supplementary Table G**.

### Pre- and peritraumatic factors are associated with the molecular recovery response

Having established the RRS as a generalizable molecular measure of recovery, we next examined whether pre-TSE clinical characteristics, prior stress burden, and acute endocrine responses were associated with RRS values (**Fig 4f and g**). Lower RRS values were consistently associated with indicators of chronic stress burden and prior adversity, including greater childhood trauma and lifetime trauma exposure, and were modestly lower among Black compared with White participants (**Fig 4f**). Conversely, higher RRS values were associated with greater psychological resilience, relational support, emotional coping capacity, and overall physical health prior to trauma exposure (**Fig 4f**).

Notably, higher peritraumatic plasma concentrations of glucocorticoids, including cortisol and other corticosteroids, were associated with higher RRS values (**Fig 4g**). None of the five medication classes examined (NSAIDs, opioids, hormonal contraceptives, benzodiazepines, and SNRIs/SSRIs) across the pre-TSE, ED, and discharge assessment periods were significantly associated with RRS values. Together, these findings suggest that greater prior stress burden is associated with an attenuated molecular recovery response, whereas stronger peritraumatic glucocorticoid responses are associated with more robust molecular recovery (**Supplementary Table 10)**.

### Recovery-associated molecular responses occur predominantly within the first two weeks following TSE

As described above, substantial molecular changes were identified between the initial ED visit and 6M post-TSE. To better understand the timing of these changes, we examined molecular data from an exploratory subset of participants (n=40) with additional blood samples collected at 2 weeks (2WK), stratified into high and low RRS groups using a median split (**Fig 5a-e**). The summed expression of the 20 RRS transcripts with higher expression at the ED decreased significantly from ED to 2WK in both groups, whereas the summed expression of the 26 transcripts with higher expression at 6M increased significantly over the same interval. In both cases, the magnitude of change in expression between ED and 2WK was greater in the high RRS group (**Fig 5a**; **Supplementary Table 11**). Overall, most transcriptional changes occurred during the ED-to-2WK interval (**Fig 5b**). Gene set enrichment analysis of ED-to-2WK changes in high RRS participants identified enrichment of myeloid cell development, homeostasis, and wound-healing pathways at the 2WK timepoint (**Fig 5c, Supplementary Table 12**).

**Fig 5.**
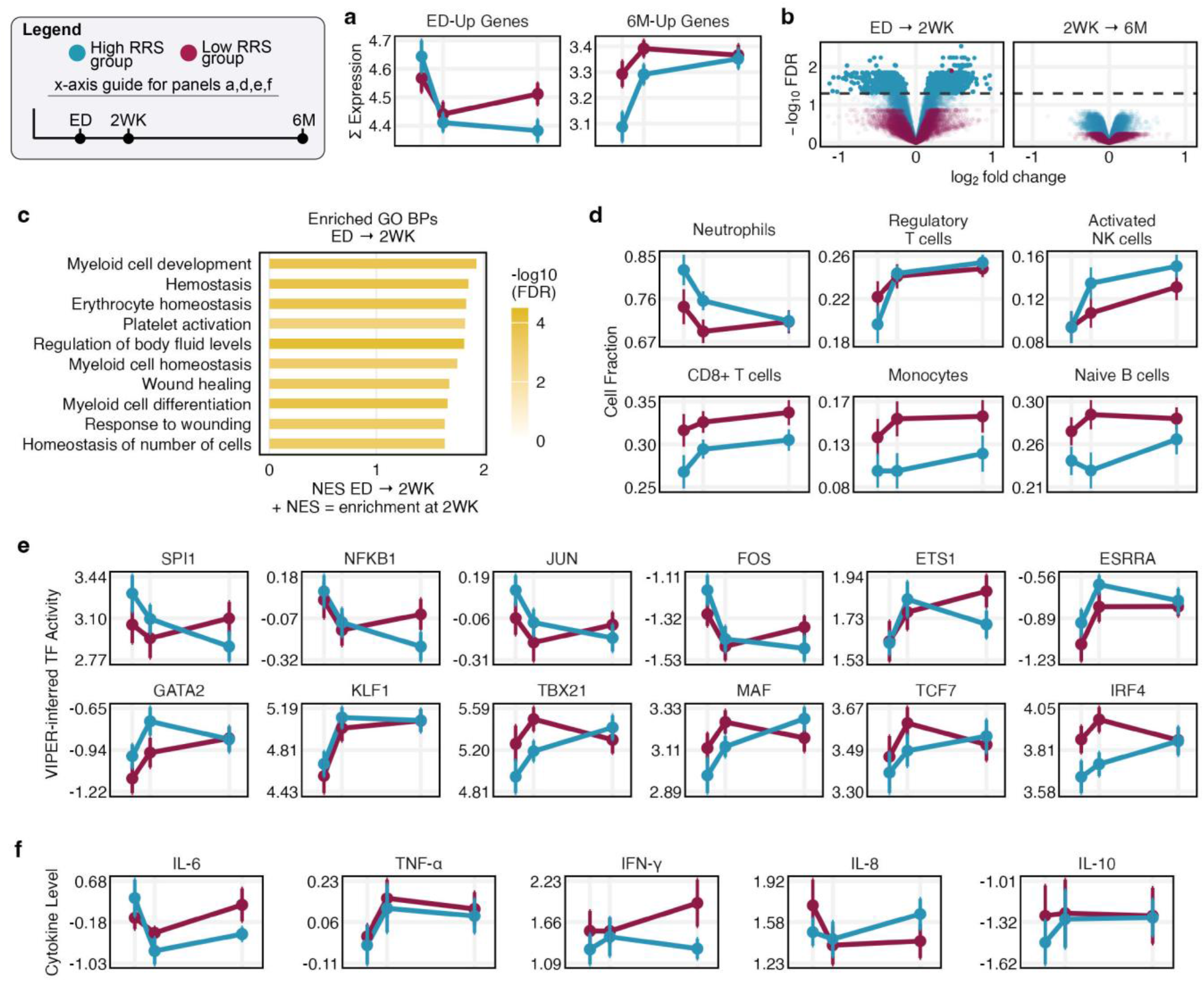
Recovery-associated molecular changes occur predominately within the first two weeks following traumatic stress exposure. Participants were stratified into high and low RRS groups using a median split. Panels a and c-f show the indicated measure across ED, 2-week (2WK), and 6-month (6M) timepoints, Panels d-f: points = mean, error bars = standard error. **a.** Summed expression of the 46 RRS genes (20 with higher expression at the ED, left; 26 with higher expression at 6M, right). **b.** Longitudinal differential expression analyses: ED to 2WK (left) and 2WK to 6M (right). Points represent genes, colored by RRS group; dashed line denotes the FDR threshold of 0.05. **c.** Enriched Gene Ontology Biological Process (GO-BP) terms from ED to 2WK differential expression analysis in high-RRS participants. **d.** Estimated immune cell fractions across timepoints. **e.** Inferred transcription factor activity for selected TFs showing significant ED-to-2WK changes. **f.** Circulating cytokine levels across timepoints; all five measured cytokines are shown.

High RRS participants also exhibited coordinated immune cell changes over this early period (**Fig 5d, Supplementary Table 13**). Estimated neutrophil fractions were highest at the ED, declined significantly by 2WK, and continued to decrease through 6M. In contrast, regulatory T cells and activated NK cells increased substantially by 2WK, whereas CD8+ T cells and monocytes increased more gradually over six months. Among low RRS participants, immune cell populations exhibited attenuated or divergent longitudinal changes compared with high RRS participants. Transcription factor activity showed a similar pattern: ETS1, GATA2, KLF1, and ESRRA increased predominantly by 2WK before plateauing in high RRS participants, whereas changes were attenuated and discordant in low RRS participants (**Fig 5e, Supplementary Table 14**).

Circulating cytokines showed similar, although not statistically significant, temporal differences (**Fig 5f, Supplementary Table 15**). High RRS participants exhibited elevated early IL-6 levels that declined by 2WK, whereas low RRS participants showed minimal early decline followed by increasing IL-6 between 2WK and 6M. TNF-α increased by 2WK among high RRS participants before plateauing, whereas IFN-γ remained stable during the first two weeks and subsequently increased through 6M among low RRS participants.

Together, these findings suggest that much of the molecular remodeling associated with recovery occurs within the first two weeks following TSE, highlighting the early posttraumatic period as a potentially important window for influencing subsequent recovery trajectories.

## Discussion

Here, we demonstrate that recovery over six months following TSE is characterized by coordinated time-dependent transcriptomic, immune cell, and cytokine changes linked to peritraumatic hormone levels. Consistent with the frequent co-occurrence of APNS (e.g., pain, posttraumatic stress, depressive symptoms) and their hypothesized shared evolutionary role in promoting survival following life threat,^27,28^ individuals who recovered exhibited remarkably similar longitudinal molecular trajectories across outcomes. As summarized in our proposed heuristic model of trauma recovery (**Fig 6**), recovery was characterized by extensive transcriptional changes, with an early neutrophil- and IL-6-associated innate immune response followed by a shift toward cellular maintenance and adaptive immune processes. In contrast, Chronic APNS trajectories exhibited comparatively limited longitudinal molecular remodeling. These findings converge with recent rodent work in which resilient animals showed a distinct hippocampal signature consistent with active molecular adaptation, suggesting that recovery across species may reflect coordinated and tissue-specific remodeling following trauma, expressed peripherally through immune and endocrine dynamics and centrally through hormone-responsive and neural-plasticity pathways.^29^ Drug-response profiling identified glucocorticoid receptor agonists and sex steroids among compounds predicted to mimic the molecular recovery response. An RRS derived from ED and 6-month transcriptomic data quantified recovery-associated molecular remodeling and generalized to an independent cohort. Higher recovery scores were associated with lower prior trauma burden and greater peritraumatic cortisol levels, while exploratory two-week data suggested that much of the recovery-associated transcriptional changes occur within this early period.

**Fig 6.**
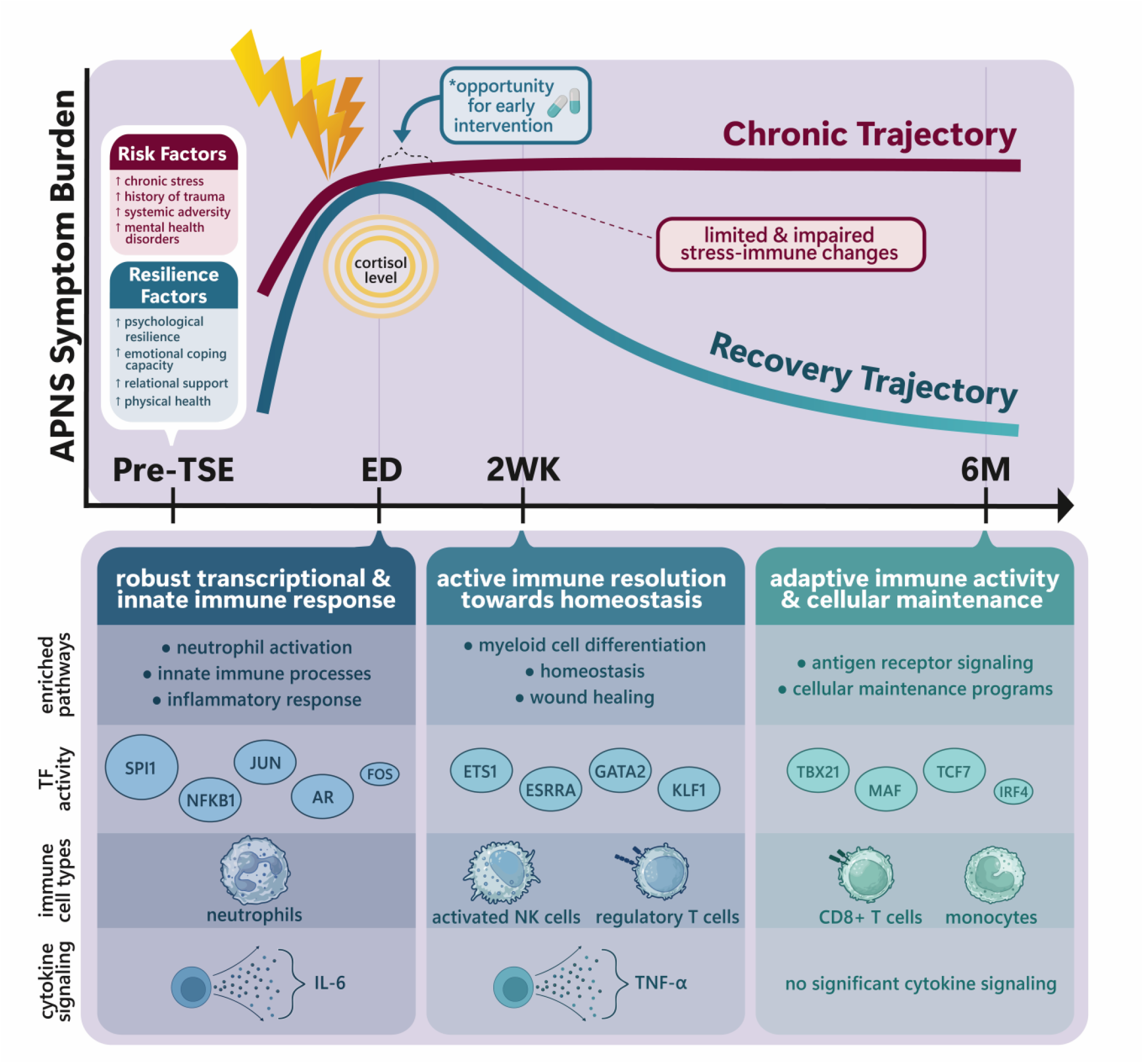
Heuristic model of the time-dependent molecular processes associated with recovery following TSE. The model integrates longitudinal transcriptomic, immune cell, transcription factor, cytokine, endocrine, and clinical findings. Individuals who recover exhibit robust early stress-immune activation at the initial emergency department (ED) visit, associated with higher peritraumatic cortisol levels and innate immune and inflammatory activation, followed by changes consistent with immune resolution and restoration of homeostasis within the first two weeks and greater adaptive immune activity and cellular maintenance by six months. Individuals with chronic APNS exhibit comparatively limited longitudinal molecular changes, potentially reflecting reduced engagement or altered coordination of recovery-associated processes. Pre-trauma risk factors (chronic stress, history of trauma and adversity, and anxiety/depression) and resilience factors (physical health, emotional coping capacity, and relational support) may contribute to this divergence. The early post-trauma period may represent an important window for preventative intervention.

As highlighted above, a central finding of these analyses is that recovery after TSE is associated with a coordinated molecular response that is evident in the immediate aftermath of TSE and evolves over time. Individuals who recovered after TSE exhibited a prominent early innate immune and inflammatory signature, including elevated neutrophil-associated transcriptional programs and circulating IL-6 levels. These early responses subsequently declined, accompanied by increasing homeostatic, tissue-repair, cellular maintenance, and adaptive immune programs over time. Together, these findings support a model in which successful recovery involves a robust early stress-immune response followed by coordinated resolution and restoration of physiological homeostasis. In contrast, Chronic trajectories were characterized by a comparatively attenuated early recovery-associated response and limited subsequent molecular remodeling, raising the possibility that persistent APNS reflect insufficient engagement and/or impaired resolution of these recovery-associated processes.

Persistent inflammatory activation is a well-established correlate of chronic pain, PTSD, depression, and other adverse posttraumatic outcomes^30,31^, and inflammation following trauma has often been framed as pathogenic.^32^ Yet, consistent with the evolutionary framework outlined above, acute innate immune activation is increasingly recognized as an adaptive, host-protective response^33^, one whose survival benefit depends on timely resolution rather than sustained activation. Emerging evidence from longitudinal human cohorts and experimental models suggests that transient early inflammatory responses may serve adaptive functions during recovery from stress and injury.^14,26,34-36^ Our findings extend this framework by demonstrating that individuals who recovered exhibited a stronger early innate immune signature than those who developed persistent APNS, followed by coordinated changes consistent with active inflammatory resolution.^37-39^ Thus, rather than supporting either sustained inflammation or broad immune suppression, our findings suggest that the timing and coordination of immune activation and resolution may be critical determinants of posttraumatic recovery.^40^

Our exploratory two-week analyses further provide insight into the timing of these recovery-associated processes. Much of the observed gene expression changes occurred within the first two weeks following TSE, with high and low RRS participants subsequently exhibiting divergent patterns of inflammatory transcription factors, cytokines, and immune cell populations. These findings suggest that recovery may depend not only on the magnitude of the early response, but also on successful transition of early stress-immune programs toward coordinated resolution. Since a substantial proportion of these gene expression changes occur within the first two weeks after trauma, this early period may represent a potentially important window during which interventions could influence long-term outcomes.^14,41-43^ Further human studies are needed to refine the timing and sequence of molecular events occurring in the initial hours, days, and weeks after trauma, and further animal-model studies are needed to test the efficacy of candidate secondary preventive interventions based on both the treatment itself and its timing of administration.^44^ Given that recovery reflects a coordinated sequence of biological processes rather than a single response, effective secondary prevention may require interventions tailored to distinct phases of the recovery process.

The observation that recovery-associated molecular trajectories diverged within the first two weeks following trauma raises an important mechanistic question: what biological systems coordinate the timing and magnitude of this early recovery program? Our findings provide convergent evidence implicating glucocorticoid signaling as a potential regulator. Multiple corticosteroid hormones measured in the ED were positively associated with molecular recovery, and glucocorticoid receptor agonists emerged from drug-response profiling as candidate modulators of the recovery-associated transcriptional program. These observations are consistent with prior studies linking blunted cortisol responses to TSE to poorer posttraumatic outcomes^45-49^ and with a broader model in which inadequate activation of the hypothalamic-pituitary-adrenal (HPA) axis contributes to posttraumatic vulnerability^50^. Although glucocorticoids are often viewed primarily as immunosuppressive hormones, they also regulate leukocyte trafficking, innate immune activation, inflammatory resolution, and transcriptional remodeling.^51,52^ Importantly, interventional studies provide additional, although mixed, support for a role of early glucocorticoid signaling in posttraumatic recovery: hydrocortisone administered shortly after TSE has reduced subsequent posttraumatic stress symptoms in some randomized trials^53-55^, whereas another randomized trial found no overall benefit.^56^ Thus, it is likely that only a subset of patients experiencing trauma would benefit from acute glucocorticoid administration, and understanding post-trauma predictive biomarkers may be critical in precision patient stratification.

In addition to glucocorticoid agonists, drug-response profiling also identified compounds predicted to induce recovery-associated transcriptional changes, including protein kinase C (PKC) activators and sex steroid hormones. These two candidate classes have some supporting evidence for potential roles in trauma recovery, including PKC activators in preclinical models^57^, and associations of peritraumatic estradiol^58,59^ and testosterone^60^ with APNS outcomes. The present study adds to this evidence, characterizing the time-dependent molecular features that distinguish recovery from APNS development and highlighting the exciting potential this holds for intervention development.

A second important translational implication of the present work is that the shared recovery program can be quantified along a continuum at the individual level. This quantification (RRS) generalized across APNS in the discovery cohort and an independent cohort. Capturing recovery through a quantitative molecular score such as the RRS, rather than through symptom-based classification alone, is a step toward closing the gap between posttraumatic outcomes whose pathogenesis is relatively well understood (e.g., wound healing) and the relative “black box” of APNS pathogenesis. Biological scores such as the RRS have several potential uses in advancing understanding of trauma-related outcomes, including identifying biological subgroups of recovery and chronicity, characterizing exposures that shape APNS vulnerability, and integrating with neuroimaging and other measures of recovery to more comprehensively characterize recovery phenotypes.

Although these findings suggest that recovery-associated biology may be therapeutically enhanced, an equally important question is why some individuals fail to exhibit a robust molecular recovery response in the first place. Individuals with lower molecular recovery had greater lifetime trauma exposure, childhood adversity, and mental health-related symptoms. Molecular recovery was also modestly lower among Black participants. Given that race is a social rather than biological construct, this association may reflect differences in cumulative trauma, adversity, structural exposures, or other social and environmental factors rather than intrinsic biological differences.^61^ Notably, low-RRS individuals did not simply show attenuated recovery-associated changes; several transcriptional, immune-cell, and transcription factor trajectories diverged or reversed direction between two weeks and six months. This pattern raises the possibility that persistent APNS may involve dysregulated temporal coordination of recovery biology rather than simply a weaker initial response.^62^ One possible explanation is that chronic stress burden alters stress-responsive endocrine systems, impairing glucocorticoid-responsive recovery programs after subsequent trauma exposure, consistent with literature implicating HPA axis dysregulation and genetic variation in *FKBP5* and *CRHR1* in APNS vulnerability.^63-66^ Future studies are needed to determine whether impaired glucocorticoid signaling represents a causal mechanism linking chronic stress burden to reduced molecular recovery.

This study has several strengths, including its longitudinal design, paired within-subject analyses, integration of transcriptomic, immune, endocrine, and cytokine data, inclusion of both men and women from diverse racial and ethnic backgrounds, and examination of ten APNS outcomes. Limitations should also be considered. Because AURORA is observational, the molecular associations identified here cannot establish whether these biological programs causally promote recovery. In addition, molecular data were available for most participants at only two timepoints (the ED and 6 months), and the sample size available for two-week analyses was modest. Immune cell fractions were estimated through computational deconvolution and transcription factor activity was inferred rather than directly measured. All molecular analyses were performed in peripheral blood, limiting inference regarding the contribution of central nervous system processes. In addition, although our findings differ from a recent AURORA analysis linking higher ED cytokines to greater PTSD severity^67^, this discrepancy likely reflects differences in analytic approach, as we examined within-person longitudinal remodeling across multiple APNS whereas Nishimi et al. modeled baseline cytokines as predictors of PTSD. Finally, although the RRS was externally validated, replication of the broader molecular findings in independent longitudinal trauma cohorts will be important.

Together, these findings provide a time-resolved molecular framework for understanding recovery following TSE. Rather than reflecting passive symptom resolution, recovery appears to involve coordinated endocrine, immune, and transcriptional programs that unfold over time and are shared across diverse posttraumatic outcomes. Although some individuals may enter the post-TSE period with biological vulnerabilities that limit engagement of these recovery-associated pathways, our findings identify recovery as an active biological process with potentially modifiable molecular components. Defining and therapeutically harnessing these endogenous recovery mechanisms may ultimately provide new opportunities to simultaneously prevent multiple APNS and reduce their substantial global burden.

## Methods

### Study design and participant cohort

#### AURORA cohort

Participants were drawn from the AURORA Study, a longitudinal observational multisite cohort study designed to investigate pain and neuropsychiatric recovery following traumatic stress exposure (TSE)^1^. Qualifying TSEs were consistent with DSM-5 Criterion A and included motor vehicle collision (MVC)s, physical or sexual assault, falls greater than 10 feet, and mass casualty incidents. Other events involving actual or threatened death, serious injury, or sexual violence were also eligible when confirmed as plausible qualifying exposures by trained research staff. Between September 2017 and July 2020, n=2,626 individuals were enrolled from 33 Emergency Departments (ED)s across the United States within 72 hours of a TSE. Participants were eligible if they had no major physical injuries (e.g., no solid organ injury Grade >1 based on the American Association for the Surgery of Trauma criteria, no significant hemorrhage, no need for a chest tube or operation with general anesthesia), were alert and oriented, fluent in English, between the ages of 18-75, and unlikely to be admitted to the hospital for >72 hours. Enrolled participants completed questionnaires and provided blood samples in the ED and were followed longitudinally for 12 months with repeated pain and neuropsychiatric assessments over that time and blood collection at two follow-up timepoints (2 weeks (2WK) and 6 months (6M)). All participants provided written informed consent under protocols approved by the institutional review boards of each participating site.

The present analysis focused on a nested longitudinal sub-cohort of 245 participants with paired whole-blood and plasma samples collected at the ED and 6M timepoints following TSE (n=490 samples). Of these participants, 40 also had blood sample available from the 2WK timepoint, enabling additional analyses of molecular changes across all three timepoints (ED, 2WK, and 6M). Demographics and clinical characteristics of the full AURORA cohort (n=2,626), the ED-to-6M nested cohort (n=245), and the ED-2WK-6M nested cohort (n=40), along with statistical comparisons across groups, are presented in Supplementary Table 1.

#### APNS trajectory classification

Symptom trajectories following TSE were previously defined and published across ten adverse post-traumatic neuropsychiatric sequelae (APNS): pain, avoidance, reexperiencing, hyperarousal, anxiety, depression, somatic symptoms, mental fatigue, nightmares and sleep disturbance^2,16^. Briefly, longitudinal symptom assessments collected via smartphone-based flash surveys administered during the post-TSE period were used to derive APNS-specific latent trajectory classes representing distinct symptom severity and recovery patterns of symptom severity and recovery over time. For each APNS, longitudinal symptom trajectories were modeled using latent growth curves, followed by growth mixture modeling to identify distinct classes based on differences in symptom intercepts and slopes. Between two and four trajectory classes were identified for each APNS, reflecting differences in symptom burden and recovery patterns following TSE.

For example, four trajectory classes were identified for pain: low (low symptom burden throughout follow-up), moderate recovery (high symptom burden in the ED with substantial recovery), moderate (high symptom burden in the ED with some recovery), and high (high symptom burden throughout follow-up). In contrast, two trajectory classes were identified for sleep disturbance: low (moderate symptom burden in the ED with substantial recovery) and high (high symptom burden throughout follow-up).

For the present analysis, trajectory classes comprising fewer than 20 participants were merged with the most phenotypically similar class within the same APNS, based on changes in symptom severity from the ED to 6 months) (calculated as Ed minus 6M symptom scores). This resulted in the merging of the following trajectory classes: the moderate recovery class with the low class for mental fatigue, and the moderate class with the high class for nightmares. For each APNS, the trajectory exhibiting the greatest reduction in symptom severity over time was designated the “Recovery” trajectory, whereas the trajectory characterized by the highest and most persistent symptom burden was designated the “Chronic” trajectory. All resulting trajectory classes (2-4 per APNS) were included in the initial longitudinal transcriptomic analyses, while downstream molecular analyses focused specifically on comparisons between the Recovery and Chronic trajectories.

#### Cross-APNS symptom comorbidity analyses

To quantify cross-APNS comorbidity, pairwise odds ratios were calculated to assess the co-occurrence of Recovery or Chronic trajectories across APNS. Odds ratios and corresponding confidence intervals were estimated from 2 x 2 contingency tables using Wald or Fisher methods, as appropriate, implemented in the *epitools^c8^* R package (v0.5.10.1). Multiple testing correction was performed using the Benjamini-Hochberg false discovery rate (FDR) procedure^69^.

#### Demographic, clinical, and medication variables

Demographic and clinical characteristics presented in Supplementary Table 1 were derived from AURORA baseline and follow-up survey data. Body mass index (BMI) was calculated from self-reported height and weight collected at the 2-week follow-up as part of the Berlin Questionnaire^70^. Sex assigned at birth was self-reported at the ED baseline visit. Age at enrollment was calculated from participants’ date of birth and ED presentation date. Educational attainment, marital status, and race/ethnicity were self-reported at the initial ED visit using standardized items from the PhenX Toolkit^71^. Peritraumatic distress was assessed at the initial ED visit using an 8-item AURORA-modified version of the Peritraumatic Distress Inventory (PDI)^72^, with item scores summed to yield a total score ranging from 0 to 32. The Area Deprivation Index (ADI), a census block group-level measure of socioeconomic disadvantage^73^, was assigned by geocoding participants’ addresses and reported as national percentile rank. Childhood trauma was assessed at the 2-week follow-up using an AURORA-modified version of the Childhood Trauma Questionnaire-Short Form (CTQ-SF)^74^ (total score range: 0-44). Lifetime traumatic event exposure was assessed at the 8-week follow-up using the Life Events Checklist for DSM-5 (LEC-5)^75^. Trauma type was classified at ED enrollment based on participant self-report and medical record review. The nested RNA-sequencing cohort included a higher proportion of participants exposed to MVCs, consistent with the study’s focus on posttraumatic recovery following events involving relatively minor physical injury. Enrollment sites were grouped into Midwest, Northeast, and Southeast geographic regions based on the location of the enrolling ED.

Pain severity during the 30 days preceding TSE and at the initial ED visit was assessed using an 11-point (0-10) Numeric Rating Scale^76^. Physical and mental health status were assessed using the SF-12 Physical and Mental Component Summary scores^77^, with norm-based scoring (population mean = 50, SD = 10). Pain interference was assessed using the PROMIS Pain Interference Short Form 4a^78^, and reported as a T-score. Anxiety was assessed using a 4-item AURORA scale adapted from items in the PROMIS Anxiety Short Form 7a^79^ and the Generalized Anxiety Disorder screener^80^, with scores reported as raw totals. Emotional capacity was assessed using the PROMIS Self-Efficacy for Managing Emotions Short Form 4a^81^ and reported as a T-score. PTSD symptom severity was assessed using the PTSD checklist for DSM-5 (PCL-5)^82^ and reported as a raw score (range 0-80); binary PTSD status was defined using a validated cutoff score greater than 30^83^. Pre-TSE depressive symptoms were assessed using the PROMIS Depression Short Form 8b^84^ and reported as a T-score; binary depression status was defined as a T-score greater than 59. Pre-TSE perceived stress was assessed using the 10-item Perceived Stress Scale (PSS-10)^85^, with scores reported as raw totals. Resilience was assessed using the 10-item Connor-Davidson Resilience Scale (CD-RISC-10)^86^ and reported as a total score.

Medication use was categorized into five classes: nonsteroidal anti-inflammatory drugs (NSAIDs), opioids, hormonal contraceptives, benzodiazepines, and serotonin-norepinephrine or selective serotonin reuptake inhibitors (SNRIs/SSRIs). For each class, binary indicators of use (yes/no) were generated for the relevant assessment periods (pre-TSE, ED presentation, and discharge prescriptions, as applicable). Medication names obtained from participant reports and medical records were mapped to World Health Organization Anatomical Therapeutic Chemical (ATC) classification codes.

### Molecular profiling and data pre-processing

#### Whole-blood and plasma collection

Whole blood was collected at the ED, 2WK, and 6M study visits using PAXgene RNA and EDTA tubes. Immediately following blood collection, PAXgene RNA tubes were mixed via inversion, incubated at room temperature for four hours, then frozen at either -20°C or -80°C at the study sites. Plasma was separated from EDTA-anticoagulated whole blood by centrifugation at 1,500 x g for 10 minutes, aliquoted into separate storage tubes, and frozen at -20°C or -80°C at the study sites. All samples were shipped in batches on dry ice to the University of North Carolina at Chapel Hill Bio-specimen Processing facility, where they were stored at -80°C until use.

#### RNA sequencing, alignment and gene quantification

Total RNA was extracted from whole-blood samples using the PAXgene Blood miRNA Kit (Qiagen, Hilden Germany) according to the manufacturer’s instructions. RNA concentration and quality were assessed using Qubit RNA Assay Kits (Thermo Fisher Scientific, Waltham, MA) and the 4200 TapeStation System (Agilent Technologies, Santa Clara, CA), respectively. Only RNA samples meeting predefined quality-control thresholds for concentration and integrity (RNA integrity number; RIN ≥ 7.0) were advanced to library preparation. Sequencing libraries were prepped from 600ng of total RNA using the Universal Plus Total RNA-Seq Library Preparation Kit with rRNA and globin depletion (TECAN, Morrisville, NC) according to the manufacturer’s specifications. Libraries were multiplexed in batches of 96, with all longitudinal samples from each participant assigned to the same sequencing batch to minimize batch-related variability in within-participant comparisons. Libraries were sequenced using 100-bp paired-end reads on an Illumina NovaSeq platform with an S4 200-cycle kit (Illumina, San Diego, CA) at the University of North Carolina at Chapel Hill High Throughput Sequencing Facility. Samples from each participant’s longitudinal timepoints were multiplexed within the same batch, precluding confounding between sequencing batch and within-participant change over time.

Raw sequencing reads were aligned to the GRCh38 human reference genome using STAR (v2.7.6a)^87^ with GENCODE v47 gene annotations. Transcript-level quantification was performed using Salmon (v1.4.0)^88^. Genes were retained for analysis if they had a mean read count of at least 5, nonzero expression in at least 90% of samples, and at least 10 reads in one or more libraries. Sample outliers were identified and removed based on principal component analysis and relative log expression quality-control metrics. Concordance between reported sex at birth and molecular sex estimates was assessed using *XIST* expression and SNP-based genetic sex, with discordant samples excluded. Following quality control, 530 RNA-sequencing samples across the three collection timepoints were retained for analysis. Transcription factor activity inference and differential expression analyses (described below) subsequently applied a gene-level expression filter of a minimum of three reads in at least one sample.

#### Inference of transcription factor activity

Transcription factor (TF) activity was inferred from whole-blood RNA-sequencing data using the *viper* package (v1.38.0)^89^ and human DoRothEA regulons^90^ (v1.16.0), which represent curated TF-target gene interactions assigned confidence levels based on the strength of supporting evidence. The *viper* algorithm infers TF regulatory activity from coordinated expression patterns among downstream target genes rather than relying solely on TF transcript abundance. Only high confidence regulons (levels A-C) containing more than 25 target genes were retained. For TF activity inference, genes with a minimum of three reads in at least one sample were retained using filterByExpr in edgeR^91^ (v4.2.2). Library sizes were normalized using trimmed mean of M values (TMM) method^92^, and normalized counts were transformed to log_2_ counts per million (CPM) using the voom-limma framework (v3.60.6)^93,94^. TF activity scores were then calculated for each sample and used to evaluate longitudinal changes in inferred TF regulatory activity.

#### Immune cell-type deconvolution

Relative immune cell-type fractions were estimated from counts per million (CPM) normalized bulk RNA-sequencing data using CIBERSORTx^95^ and the LM22 leukocyte signature matrix, which contains reference expression profiles for 22 immune cell subsets. Cell types with a median estimated fraction of zero across the cohort were excluded, yielding 14 consistently detected immune cell populations for downstream analyses: neutrophils, resting mast cells, activated dendritic cells, M2 macrophages, monocytes, activated NK cells, resting NK cells, regulatory T cells, CD8+ T cells, resting memory CD4+ T cells, naive CD4+ T cells, plasma cells, memory B cells, and naive B cells. Estimated cell-type fractions were arcsine square root-transformed to stabilize variance and improve approximation to normality for downstream statistical analyses.

#### Cytokine quantification and data preprocessing

Cytokines (IL-10, IL-6, IL-8, TNF-α, and IFN-γ) were measured in multiplex as previously described^96^. Briefly, plasma samples were diluted and quantified using Meso Scale Discovery (MSD)’s V-PLEX Proinflammatory Panel 1 Human chemiluminescence-based assay (Gaithersburg, MD, USA, catalog # K15049D-1) based on manufacturer’s specifications at The University of North Carolina at Chapel Hill. Assay panel plates were read and analyzed immediately following preparation using a QuickPlex SQ 120 instrument (MSD) and the DISCOVERY WORKBENCH® 4.0 software. The intra- and inter-assay coefficient of variation between samples and plates was ≤ 15%. The few samples with higher coefficients of variation were repeated or excluded from the analysis.

Cytokine concentrations were log-transformed before analysis. Potential plate and batch effects were evaluated separately for each cytokine using linear models that included assay plate, batch, and collection timepoint, allowing technical effects to be evaluated while accounting for biological changes over time. Significant batch effects were observed for TNF-α and IL-8, whereas plate effects were observed across all cytokines. To preserve longitudinal biological variation, only technical effects (plate and/or batch), and not collection timepoint, were removed during adjustment. Log-transformed TNF-α and IL-8 values were adjusted for both plate and batch, while IL-10, IL-6, and IFN-γ values were adjusted for plate only. Adjusted cytokine values were calculated as the log-transformed concentrations minus the estimated contributions of the relevant technical effects, preserving variation associated with collection timepoint.

For longitudinal analyses, cytokine data were restricted to participants in the RNA-sequencing cohorts with complete measurements at the relevant timepoints. ED-to-6M analyses included participants with cytokines measurements at both ED and 6M, while ED-2WK-6M analyses included participants with measurements at all three timepoints.

#### Hormone measurement

Plasma samples were shipped on dry ice to Biocrates Life Sciences (Innsbruck, Austria) for quantification of seventeen steroid hormones including cortisol, corticosterone, and dehydroepiandrosterone DHEA), using the AbsoluteIDQ® Stero17 assay. Hormones were quantified using ultra-performance liquid chromatography–tandem mass spectrometry (UPLC-MS/MS) in multiple reaction monitoring mode on a Waters XEVO TQ-S micro instrument (Waters, Vienna, Austria). Prior to analysis, samples underwent solid-phase extraction (SPE) in a 96-well plate format to isolate and concentrate steroid hormones. Plasma samples, calibrators, and quality control samples were spiked with internal standards and applied to a preconditioned SPE plate. After drying, analytes were eluted twice with dichloromethane into a capture plate and subsequently evaporated under nitrogen gas. Samples were then reconstituted in 25% methanol in water for UPLC-MS/MS analysis. Quantification was performed using seven-point external calibration curves and 15 stable isotope-labeled internal standards. Quality control samples at three concentration levels were included to assess assay performance. Steroid hormone concentrations were calculated using Biocrates’ WebIDQ software^97^.

Samples identified as hemolytic or turbid were excluded. Hormone analyses focused on ED plasma concentrations among 184 participants with paired ED-6M RNA-sequencing data. Values below the analyte-specific limit of detection (LOD) were imputed as LOD/√2. Hormone concentrations were subsequently log_2_ transformed to reduce right skewness. Potential variation associated with sample preparation plates was evaluated separately for each hormone using linear models, and no significant plate effects were detected.

### Longitudinal analysis of molecular recovery

Longitudinal analyses were performed using paired within-participant designs to quantify molecular changes between the ED and 6M timepoints. Analyses were performed separately for each trajectory across the ten APNS.

Longitudinal differential expression analyses were performed using the *dream* framework from the *variancePartition* package (v1.34.0)^98^, in conjunction with the *voom-limma* framework (v 3.60.6)^93,94^, with participant included as a random effect to account for repeated measures. Gene-level read counts were filtered to retain genes with a minimum of three reads in at least one sample. Library sizes were normalized using the trimmed mean of M-values (TMM) method, and expression values were transformed to log_2_ counts per million (CPM) using *voom*. The *dream* framework incorporates *voom*-derived observational precision weights to account for the mean-variance relationship inherent to RNA-sequencing data. Differential expression statistics were subsequently moderated using empirical Bayes methods^99^. Because all longitudinal samples from a given participant were sequenced within the same batch, the paired design also minimized potential confounding from between-batch technical variation.

To assess whether differences in longitudinal differential expression were attributable to trajectory sample size, sensitivity analyses were conducted using repeated random downsampling. For each trajectory across all APNS, 20 participants with paired ED and 6M samples were randomly selected without replacement, and differential expression was reassessed using the same filtered, TMM-normalized gene set and *dream* analysis pipeline described above. This procedure was repeated for 10 independent iterations per APNS trajectory, and the median number of differentially expressed genes with FDR<0.05 across iterations was used to summarize the longitudinal transcriptional signature for each trajectory.

For inferred TF activity, estimated immune cell fractions, and circulating cytokine measurements, within-individual longitudinal change scores were computed as Δ = 6M − ED. Longitudinal changes within each APNS trajectory were evaluated using two-sided paired T-tests. P-values were adjusted for multiple comparisons within each molecular modality using the Benjamini-Hochberg procedure.

### Gene set enrichment analyses

Gene set enrichment analyses were performed using the FGSEA algorithm (v1.30.0)^100^ to identify biological pathways associated with longitudinal transcriptional changes associated with clinical recovery following TSE across all APNS. For each APNS, gene-level statistics were obtained from paired differential expression analyses comparing ED and 6M timepoints within the Recovery trajectory. For each APNS, gene-level statistics were obtained from paired differential expression analyses comparing ED and 6M timepoints within the Recovery trajectory. Genes were ranked by a weighted score reflecting the magnitude, direction of effect, and significance calculated as log₂ fold change × −log₁₀(p-value). Gene set enrichment analyses were performed on the single ranked list for each APNS, with pathways enriched at the ED visit indicated by a negative normalized enrichment score (NES) and 6M enrichment pathways represented by a positive NES. Enrichment analyses were performed using two pathway collections: Gene Ontology Biological Process (GO:BP) and Reactome pathways, both obtained from the Molecular Signatures Database (MSigDB; v7.5.1; C5:GO:BP and C2:CP:REACTOME, respectively)^101^. Gene sets containing 50-500 genes were included in the analyses. Significant pathway enrichment was defined as a Benjamini-Hochberg FDR<0.01.

### Associations between transcriptional and immune cell changes

Associations between longitudinal changes in gene expression and estimated circulating immune cell fractions were evaluated using within-participant change scores (Δ = 6M − ED). Gene expression changes were calculated for genes exhibiting concordant differential expression across the Recovery trajectories of all ten APNS, and corresponding changes in estimated immune cell fractions were calculated for each participant over the same interval.

For each immune cell population, separate univariable linear regression models were fitted for each gene to assess associations between changes in gene expression and changes in estimated immune cell fractions. Associations were evaluated using two-sided tests, and p-values were adjusted for multiple testing using the Benjamini-Hochberg procedure separately across genes for each immune cell population.

### Pharmacologic perturbation analyses

Connectivity Map (CMap) analyses were performed using the LINCS CLUE platform^17,102^ to identify pharmacologic perturbations whose transcriptional signatures were concordant or discordant with recovery-associated gene-expression changes. CMap is a large-scale reference resource comprising gene expression signatures induced by small-molecule and genetic perturbations across diverse human cell lines. Transcriptional profiling in LINCS is performed using the L1000 platform, which measures expression of approximately 1,000 landmark genes and computationally infers the expression of additional genes, enabling high-throughput screening of perturbational responses^103^. Three recovery-associated query signatures were generated from ED-to-6M longitudinal differential expression analyses using the top 50, 75, and 100 upregulated and downregulated genes exhibiting concordant differential expression across the Recovery trajectories of all ten APNS overlapping the LINCS BING space. Each signature was submitted independently to CLUE, and the corresponding gene lists are provided in **Ext. Data Fig 5**.

CLUE query results were annotated with cell-line metadata and restricted to small-molecule perturbations. Signatures from central nervous system, autonomic ganglia, placenta, and unknown cell lineages were excluded. Downstream analyses retained signatures meeting LINCS quality-control criteria, including CLUE high-confidence connectivity designation and a transcriptional activity score (TAS) ≥ 0.2. Positive normalized connectivity scores (NCS) were interpreted as indicating transcriptional concordance with recovery-associated signatures (recovery mimickers), whereas negative NCS values indicated opposing transcriptional patterns (recovery opposers). Signatures meeting these criteria were considered qualifying signatures for downstream compound-level analysis.

For compound-level aggregation, perturbation signatures were stratified by NCS direction and summarized separately as mimickers or opposers. Compounds were required to have at least four qualifying signatures in the corresponding direction. Compound-level summaries included median NCS, NCS variability, median TAS, number of qualifying signatures, number of cell lines, annotated mechanisms of action, and predicted targets.

### Derivation of the RNA-based Recovery Score (RRS)

#### Extreme phenotype definition

To define extreme recovery phenotypes across APNS, the number of Recovery and Chronic trajectory assignments was calculated for each participant across the ten APNS. Because Recovery and Chronic trajectory assignments differed in their frequency and patterns of co-occurrence across APNS (**Ext. Data Fig 3**), asymmetric thresholds were evaluated to identify distinct extreme phenotypes while preserving adequate sample sizes. Multiple combinations of Recovery and Chronic APNS trajectory thresholds were evaluated to assess group size and overlap between extreme phenotype groups (**Ext. Data Fig 6a**). Thresholds were selected based on clinical trajectory distributions and group sizes, independently of gene-expression data.

Participants assigned to Recovery trajectories in at least seven APNS and Chronic trajectories in fewer than two APNS were classified as “Most-Recovered” (n = 96). Participants assigned to Chronic trajectories in at least two APNS and Recovery trajectories in fewer than seven APNS were classified as “Most-Chronic” (n = 82). These thresholds eliminated overlap between the Most-Recovered and Most-Chronic groups while retaining sufficient participants for model development and evaluation. All remaining participants were classified as “Intermediate” (n = 67) and were excluded from model training and feature selection.

#### RRS derivation

Participant-level longitudinal transcriptional changes were quantified as ED-to-6M log_2_ fold-changes for recovery-associated genes expressed across all ten APNS Recovery trajectories. These longitudinal gene expression changes served as input features for development of the RNA-based Recovery Score (RRS), a quantitative measure of recovery-associated transcriptional remodeling following TSE.

Elastic net regularized logistic regression was performed to identify longitudinal transcriptional features distinguishing Most-Recovered and Most-Chronic phenotypes. Analyses were restricted to the extreme phenotype cohort, which was randomly partitioned into stratified training (80%) and independent heldout test (20%) cohorts, preserving the balance between Most-Recovered and Most-Chronic participants. Intermediate participants were retained as an additional heldout group and were not included in model fitting or feature selection. Sex and race distributions were compared across the AURORA training, heldout test, and Intermediate cohorts to assess demographic comparability (**Ext. Data Fig 6b**).

Within the training cohort, repeated Monte Carlo subsampling with bootstrap-based stability selection was performed. Across 20 Monte Carlo outer splits, a stratified random 80% subsample of the training cohort was selected for model fitting. Within each outer split, 50 balanced bootstrap samples were generated by resampling equal numbers of Most-Recovered and Most-Chronic participants with replacement. For each bootstrap iteration, elastic net models were fit using the glmnet package (v4.1-8) with α = 0.1 and λ selected using 10-fold cross-validation via the one-standard-error rule, favoring a more parsimonious model over the model with the minimum cross-validation error. Genes with nonzero coefficients were retained from each bootstrap model.

Gene stability was evaluated across bootstrap iterations and Monte Carlo outer splits. Threshold sensitivity analyses were performed to evaluate the effect of varying within-split and outer-split stability thresholds on final signature size. Lower thresholds produced substantially larger gene sets, whereas increasingly stringent thresholds rapidly reduced the number of retained genes (**Ext. Data Fig 6c**). Final thresholds of ≥50% within-split stability and ≥40% outer-split stability were selected to balance signature size and feature stability, retaining sufficient genes to capture interindividual variability and facilitate overlap with external transcriptomic datasets while limiting inclusion of unstable features.

For each gene retained following stability selection, elastic net coefficients were first averaged across the 50 bootstrap models within each Monte Carlo outer split. The resulting split-specific coefficients were then averaged across the 20 Monte Carlo splits to generate the final weight for each gene (**Ext. Data Fig 6d**). Participant-level RRS values were calculated as the weighted sum of each participant’s longitudinal ED-to-6M log2 fold-changes using these final gene weights. The same gene weights were applied without modification to the training cohort, heldout test cohort, Intermediate participants, and external validation cohort.

#### Internal and external evaluation

RRS performance was evaluated in both the training and independent heldout cohorts by comparing Most-Recovered and Most-Chronic participants using two-sided Welch’s t-tests. Discrimination between these phenotypes was additionally assessed using receiver operating characteristic (ROC) curve analysis and the area under the curve (AUC).

To evaluate associations between the RRS and individual APNS outcomes, RRS values were merged with latent trajectory classifications for each APNS. For each APNS, participants assigned to Chronic trajectories were compared with those assigned to all other trajectories using ROC analyses and AUC estimates. Associations between RRS and APNS latent trajectory classifications were additionally evaluated using Welch’s t-tests for binary outcomes and one-way ANOVA for APNS with more than two trajectory classes. For APNS with more than two trajectories, post-hoc pairwise comparisons were performed using estimated marginal means from the *emmeans* package (v1.11.1). P-values were adjusted for multiple testing across APNS using the Benjamini-Hochberg FDR procedure.

External validation was performed using an independent longitudinal whole-blood RNA-sequencing cohort of individuals with acute low back pain who provided blood samples at an early assessment and approximately six months later (PainOMICs, GEO accession GSE177034). Raw count data were imported, mapped from Entrez gene identifiers to HGNC gene symbols, and log_2_ transformed after addition of a pseudocount. Participants with paired baseline (t0) and follow-up (t1) RNA-sequencing samples were retained for analysis.

The final gene weights derived from the AURORA training cohort were applied directly to the external cohort without retraining or coefficient modification. Participant-level longitudinal transcriptional changes were calculated as log_2_ expression differences between t1 and t0 for genes overlapping with the final RRS signature. These transcriptional changes were multiplied by their corresponding gene weights and summed to calculate participant-level RRS values. Associations between RRS values and persistent versus resolved pain outcomes were evaluated using two-sided Welch’s t-tests.

### Correlates of the RNA-based Recovery Score

To identify participant characteristics associated with the RRS, univariate linear regression analyses were performed across four pre-specified categories of predictors: (1) demographic and baseline physical health measures, (2) baseline mental health, stress and adversity exposures, (3) pre- and peritraumatic medication use, and (4) peritraumatic steroid hormone levels measured in plasma collected at the ED visit. Definitions, measurement scales, and coding procedures for all evaluated variables are provided in **Supplementary Table 10**.

Separate linear regression models were fitted for each predictor, with RRS as the outcome. Continuous predictors were standardized to a mean of zero and standard deviation of one before analysis, allowing regression coefficients (β) to represent the difference in RRS associated with a one-standard-deviation increase in the predictor. Regression coefficients were reported with corresponding 95% confidence intervals, and p-values were adjusted for multiple testing within each predictor category using the Benjamini-Hochberg FDR procedure.

### Acute and delayed longitudinal molecular changes

Additional longitudinal analyses were conducted in the subset of 40 participants with available whole-blood samples collected at 2WK following TSE, in addition to paired ED and 6M samples. Participants were stratified into high and low RRS groups using the median RRS value from the full cohort of 245 participants as the cutoff. Longitudinal molecular changes were evaluated across the ED-to-2WK, 2WK-to-6M, and ED-to-6M intervals using gene-expression profiles, inferred TF activity, estimated immune cell fractions, and circulating cytokine measurements.

Within-group longitudinal changes across each interval were evaluated using two-sided paired t-tests within the high- and low-RRS groups. Each molecular modality was processed and analyzed using the corresponding preprocessing and longitudinal analysis procedures described above. Gene set enrichment analyses for the 2WK subset were performed using the same FGSEA workflow, pathway collections, gene-set size filters, and gene-ranking approach described above. These enrichment analyses were restricted to the high-RRS group and evaluated the ED-to-2WK and ED-to-6M intervals, in which differential expression was observed. For these analyses, pathways with Benjamini–Hochberg FDR<0.05 were considered significant.

## Data availability

The datasets used in this manuscript are available at the following link: https://doi.org/10.15139/S3/XSY1ZR (until publication, the content can be viewed at the following preview URL: https://dataverse.unc.edu/previewurl.xhtml?token=05ff18fc-df69-4798-8aaf-306f3eda517b).

## Code availability

All code used for this study is publicly available on GitHub at https://github.com/brittaniewinfield-UNC/aurora-trauma-recovery-rnaseq.

## Acknowledgments

The investigators wish to thank the trauma survivors participating in the AURORA Study. Their time and effort during a challenging period of their lives make our efforts to improve recovery for future trauma survivors possible. The AURORA study was supported by NIMH under U01MH110925, the US Army MRMC, One Mind, and The Mayday Fund. The DISENTANGLE study is a continuation of AURORA’s work and is supported by the United States Army Medical Research Acquisition Activity (USAMRAA) under Contract No. W81XWH22C012. Mindstrong Health provided some of the software used to perform study assessments. Research reported in this publication was additionally supported by NIAMS, NINDS, and NIGMS of the National Institutes of Health under Award numbers, R01AR081454 (Linnstaedt), R01NS118563 (Linnstaedt and McLean), and (5T32GM135123 (Winfield). Additional funding was provided by the Rita Allen Foundation (Linnstaedt) and the Yang Family Biomedical Scholars Award (Linnstaedt). Data used in this manuscript is available through the National Institute of Mental Health (NIMH) Data Archive (NDA). The NDA Collection for the AURORA Project can be found here: https://nda.nih.gov/edit_collection.html?id=2526. This content is solely the responsibility of the authors and may not reflect the official view of any of the funders or the Submitters submitting original data to NDA.

---

**Ext Data Fig 1.**
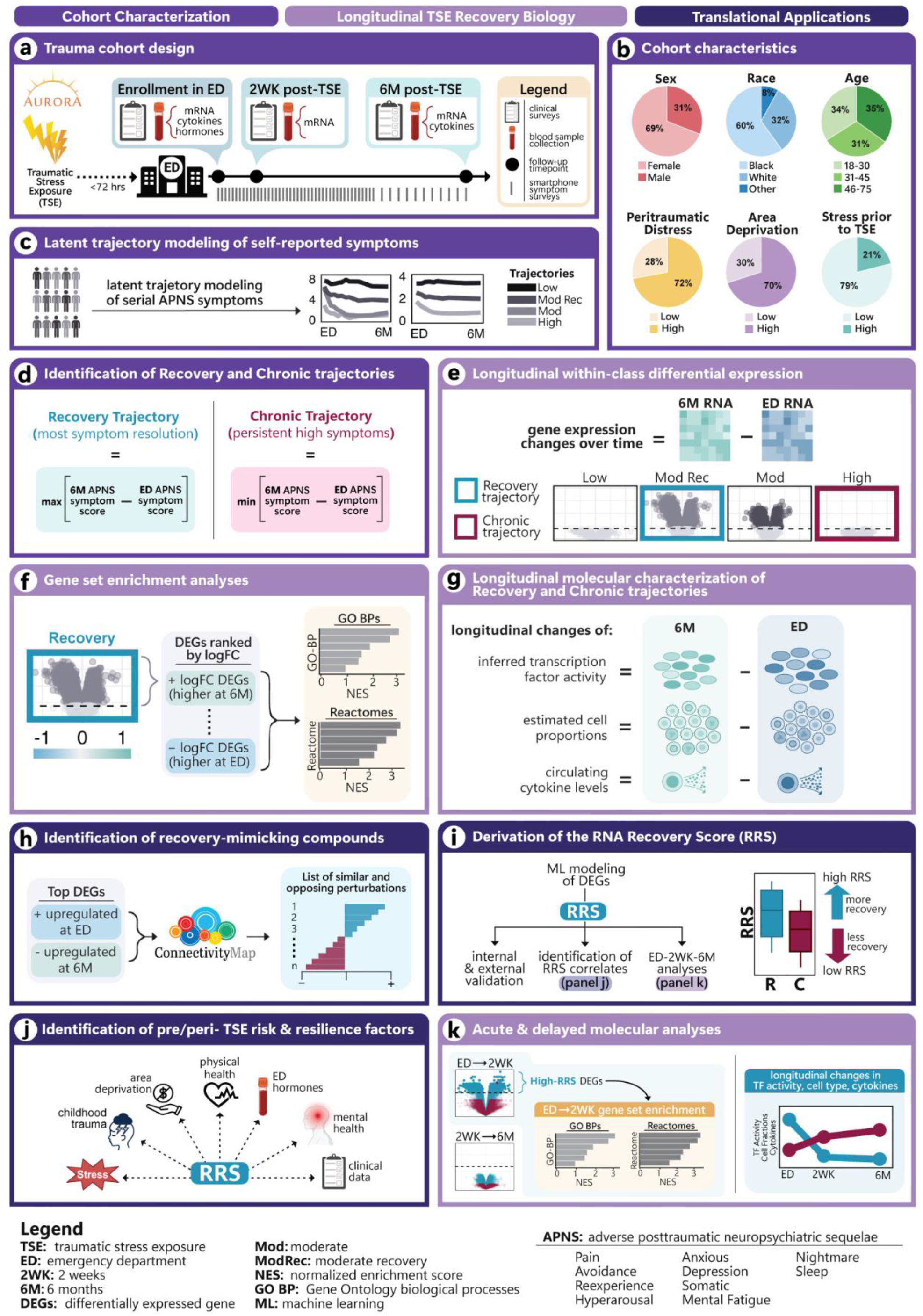
Study design and analytic workflow for characterizing molecular features of recovery after traumatic stress exposure. **a.** Longitudinal design of the AURORA cohort and blood collection timepoints for the current nested study. **b.** Demographic and selected clinical characteristics of study participants (Supplementary Table 1). **c.** Latent trajectory class modeling of longitudinal adverse posttraumatic neuropsychiatric sequelae (APNS) symptom severity, yielding 2-4 trajectories per APNS, as described previously^2,16^. **d.** Identification of the Recovery and Chronic trajectories across each APNS, representing the greatest symptom resolution and most persistent symptoms, respectively. **e.** Longitudinal, paired differential expression analyses for all trajectories across the ten APNS. (Results, Fig. 1; Supplementary Table 2). **f.** Gene set enrichment analyses for the Recovery trajectories across each APNS. Genes ranked by differential expression statistics were assessed for enrichment of Gene Ontology Biological Processes (GO-BP) terms and Reactome pathways. (Results, Fig. 2b; Supplementary Table 4). **g.** Longitudinal, paired analyses across APNS trajectories to identify time-dependent changes in inferred transcription factor activity, estimated immune cell fractions, and circulating cytokine levels. (Results, Fig. 2c-e; Supplementary Tables 5-7). **h.** Identification of compounds predicted to mimic or oppose recovery-associated transcriptional patterns using recovery-associated differentially expressed genes and the Connectivity Map (LINCS L1000) (Results, Fig. 3; Supplementary Table 8). **i.** Generation of an RNA-based Recovery Score (RRS) to quantify recovery-associated molecular changes. The RRS was evaluated in a held-out AURORA cohort and externally validated in PainOMICs, then used to examine pre- and peri-TSE risk factors (panel j) and the timing of recovery-associated molecular changes (panel k). (Results, Fig. 4). **j.** Identification of pre- and peri-TSE factors associated with molecular recovery using the RRS and demographic, clinical, psychosocial, and ED hormone measurements. (Results, Fig. 4f,g; Supplementary Table 10). **k.** Exploratory characterization of the timing of recovery-associated molecular changes using the RRS, RNA expression profiles, estimated immune cell fractions, inferred TF activity, and circulating cytokines across the three blood collection timepoints in AURORA: ED, 2 weeks, and 6 months. (Results, Fig. 5; Supplementary Tables 11-15)

**Ext Data Fig 2.**
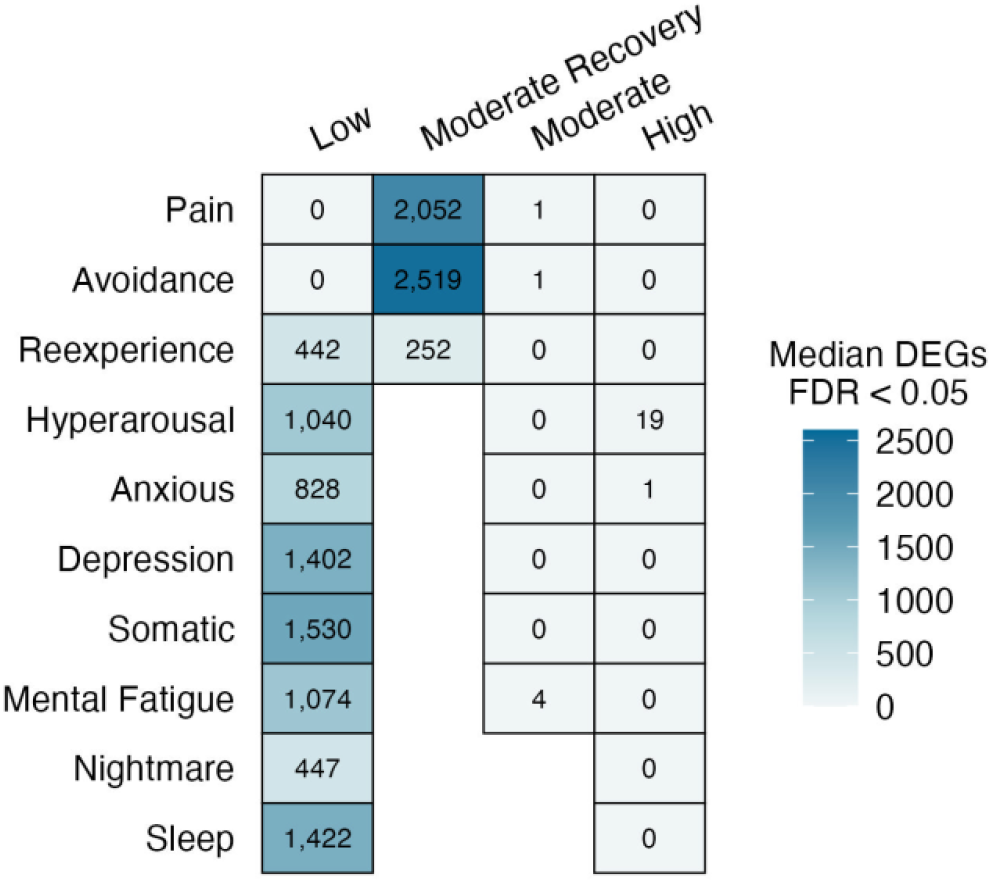
Sensitivity analysis of longitudinal differential expression following equal-sized downsampling across APNS trajectories. Median number of differentially expressed genes (DEGs, FDR<0.05) across 10 iterations of random downsampling to 20 paired participants per APNS trajectory, performed to assess whether differences in DEG counts identified in the primary longitudinal analysis (Fig. 1) reflected differences in trajectory sample size rather than underlying transcriptional changes. For each trajectory across the ten APNS, differential expression between the ED and 6-month timepoints was reassessed in 10 independently downsampled subsets of 20 paired participants using the same filtered, trimmed mean of M-values (TMM)-normalized gene set and voom-limma pipeline as the primary analysis; cell values indicate the median DEG count across iterations. Cells are shaded by median DEG count, with dark blue representing a greater number of DEGs. Blank cells indicate APNS domains for which a corresponding trajectory class was not identified.

**Ext Data Fig 3.**
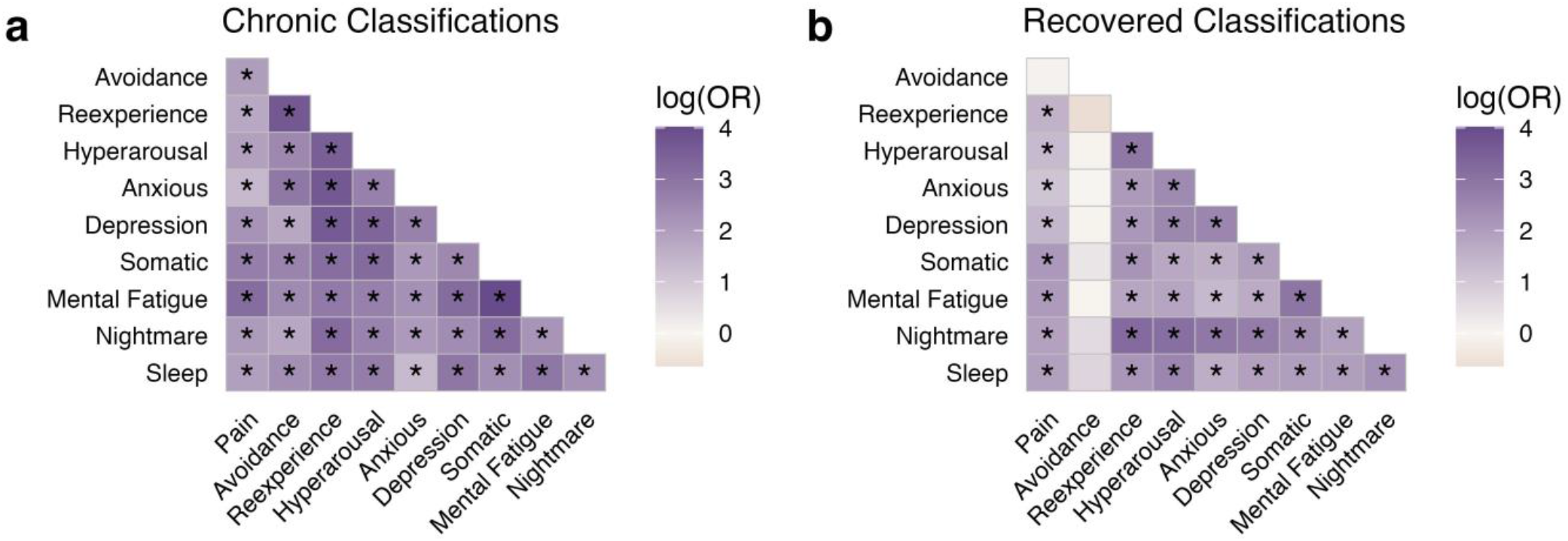
Cross-APNS co-occurrence of Chronic and Recovery trajectory classifications. Heatmaps show pairwise log odds ratios (log(OR)) for the co-occurrence of **a.** Chronic and **b.** Recovery trajectory classifications across the ten APNS. Odds ratios were calculated from 2x2 contingency tables using Wald tests or Fisher’s exact tests, as appropriate. Darker purple shading indicates higher log(OR) values, reflecting stronger associations between trajectory classifications across APNS pairs; near white or pink shading indicates a weak or negative association. Asterisks indicate statistically significant associations (FDR<0.01).

**Ext Data Fig 4.**
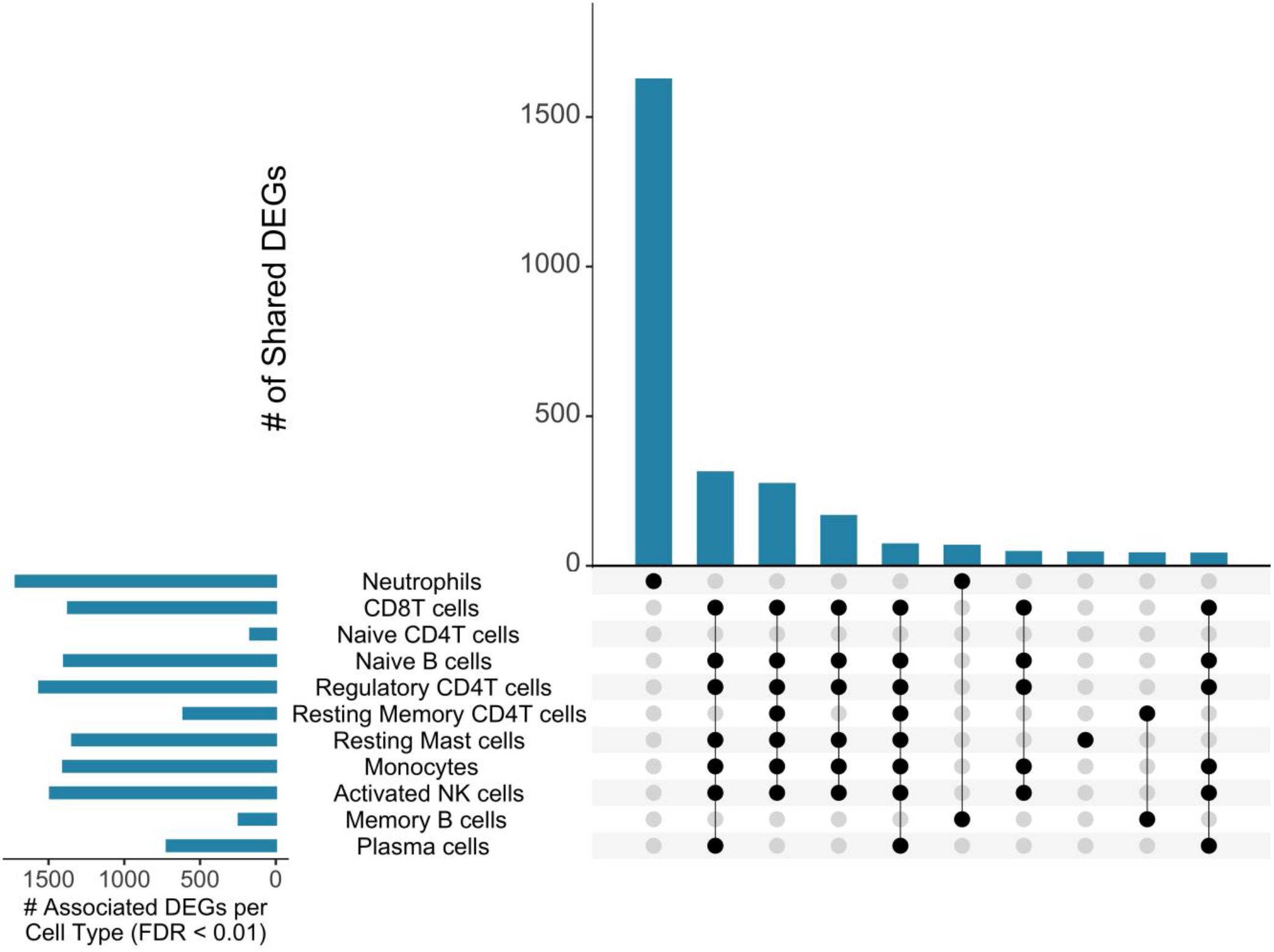
Relationship between shared recovery-associated differentially expressed genes and estimated immune cell-type fractions. Associations between longitudinal transcriptional changes and changes in circulating immune cell fractions were evaluated using within-participant change scores (Δ = 6M − ED) for the 3,574 shared recovery-associated differentially expressed genes (DEGs) identified across the Recovery trajectories for all ten APNS and for estimated immune cell-type fractions derived from CIBERSORTx deconvolution. For each immune cell population, linear regression models were fit independently for each gene to assess the association between longitudinal changes in gene expression and changes in immune cell fractions; significance was evaluated using two-sided tests, with Benjamini-Hochberg FDR correction applied separately within each cell population. The UpSet plot displays intersections of DEGs significantly positively associated (FDR < 0.01) with changes in estimated immune cell fractions across eleven cell types, showing the ten largest intersections. Horizontal bars (bottom left) indicate the total number of DEGs positively associated with each cell type; vertical bars (top) indicate the number of DEGs in each intersection, as defined by the connected black dots below each bar.

**Ext Data Fig 5.**
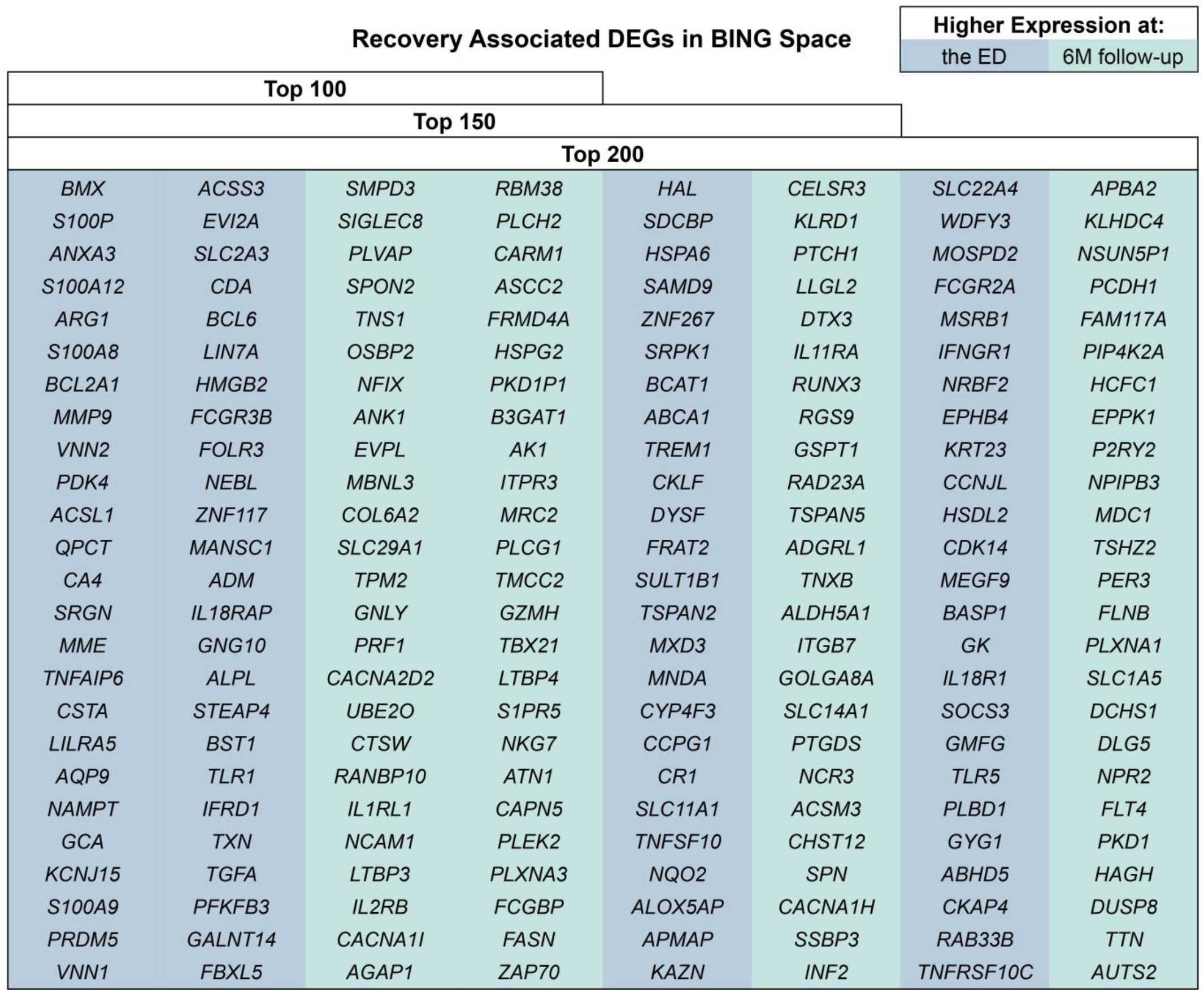
Recovery-associated differentially expressed genes used in Connectivity Map analyses. Recovery-associated differentially expressed genes, shared across the Recovery trajectories for all ten APNS and ranked by average absolute log2 fold-change from the ED-to-6M longitudinal differential expression analysis, were restricted to genes represented in the LINCS BING (Best Inferred Genes) space and submitted as Connectivity Map (CMap) query gene sets via the LINCS CLUE platform (Fig. 3). Query sets combined the top 50, 75, or 100 genes with higher expression at 6M with the top 50, 75, or 100 genes with higher expression at the ED, yielding nested Top 100, Top 150, and Top 200 gene sets submitted as independent CLUE queries. Genes shaded blue exhibited higher expression at the emergency department timepoint relative to the 6M follow-up; genes shaded teal exhibited higher expression at the 6M timepoint relative to the ED.

**Ext Data Fig 6.**
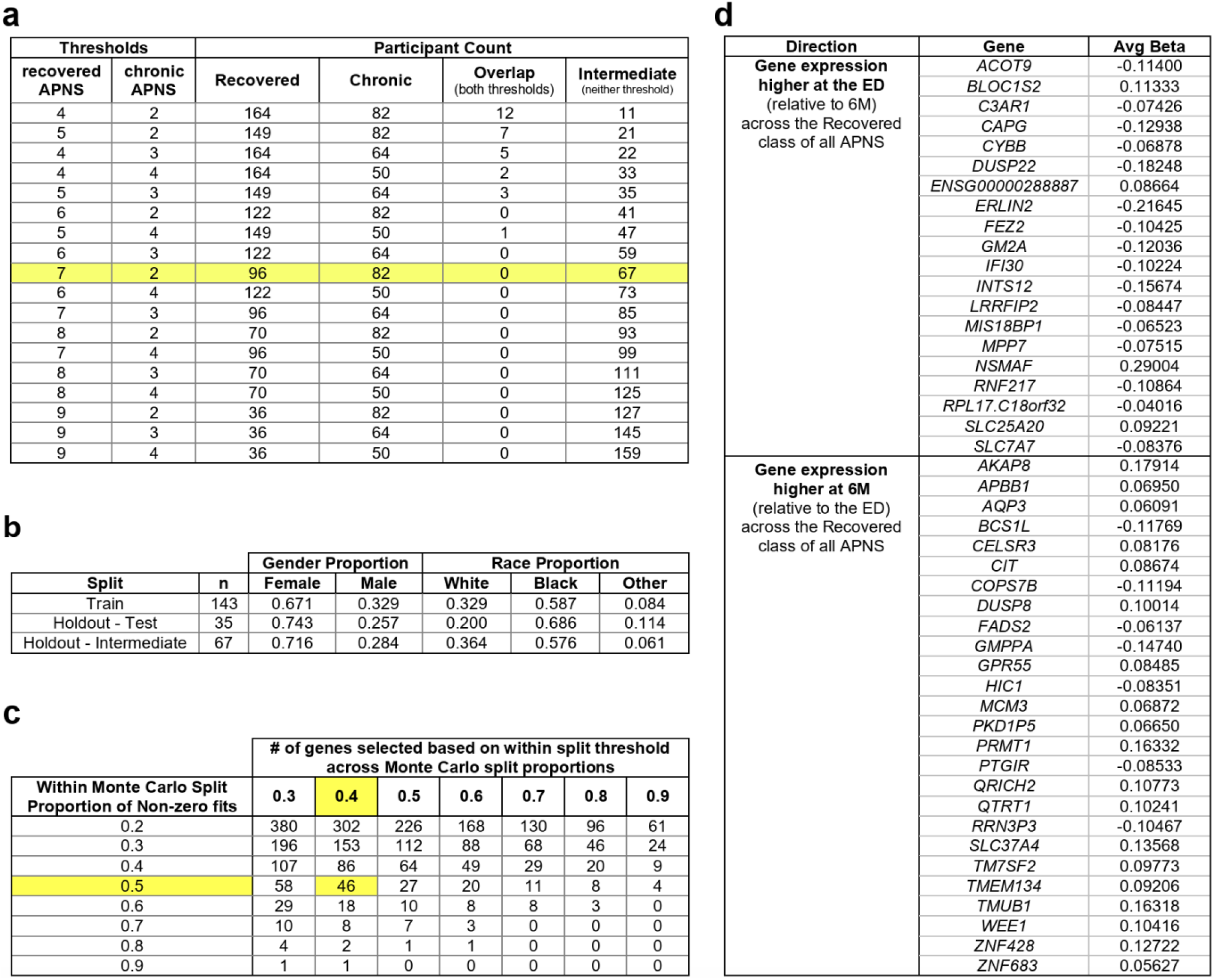
Derivation of the RNA-based Recovery Score. RNA-Based Recovery Score (RRS) derivation used longitudinal ED-to-6M log2 fold-changes in recovery associated differentially expressed genes shared across the ten Recovery APNS trajectories as input features for elastic net regularized logistic regression to distinguish Most-Recovered and Most-Chronic phenotypes. **a.** Participant counts across combinations of recovery and chronic APNS thresholds evaluated to define extreme recovery phenotypes. The thresholds used to define Most-Recovered (≥7 Recovery trajectories, <2 Chronic trajectories) and Most-Chronic (≥2 Chronic trajectories, <7 Recovered trajectories) groups are highlighted; remaining participants were classified as Intermediate. **b.** Sex and race proportions across the training, held-out test, and Intermediate cohorts used for model development and evaluation. **c.** Number of genes retained following bootstrap-based stability selection across a range of within-split and across-split stability thresholds, evaluated across 20 Monte Carlo outer splits and 50 balanced bootstrap samples per split. The thresholds used for the final signature (≥50% within-split stability, ≥40% across-split stability) are highlighted. **d.** Final elastic net gene weights averaged first across bootstrap models within each Monte Carlo outer split and then across all outer splits, for genes with higher expression at the ED relative to 6M (top) and higher expression at 6M relative to ED (bottom). These weights were applied without modification to calculate participant-level RRS values in the training, held-out, Intermediate, and external validation cohorts.

